# Moment-to-moment brain signal variability reliably predicts psychiatric treatment outcome

**DOI:** 10.1101/2021.02.17.21251814

**Authors:** Kristoffer N. T. Månsson, Leonhard Waschke, Amirhossain Manzouri, Tomas Furmark, Håkan Fischer, Douglas D. Garrett

## Abstract

Biomarkers of psychiatric treatment response remain elusive. Functional magnetic resonance imaging (fMRI) has shown promise, but low reliability has limited the utility of typical fMRI measures as harbingers of treatment success. Strikingly, temporal variability in brain signals has already proven a sensitive and reliable indicator of individual differences, but has not yet been examined in relation to psychiatric treatment outcomes. Here, 45 patients with social anxiety disorder were scanned twice (11 weeks apart) using simple task-based and resting-state fMRI to capture moment-to-moment neural variability. After fMRI test-retest, patients underwent a 9-week cognitive-behavioral therapy. Reliability-based 5-fold cross-validation showed that task-based brain signal variability was the strongest contributor in a treatment outcome prediction model (total *r*_ACTUAL,PREDICTED_ = .77) - outperforming self-reports, resting-state neural variability, and standard mean-based measures of neural activity. Notably, task-based brain signal variability showed excellent test-retest reliability (intraclass correlation coefficient = .80), even with a task length less than 3 minutes long. Rather than a source of undesirable “noise”, moment-to-moment fMRI variability may instead serve as a highly reliable and efficient prognostic indicator of clinical outcome.

---

Biomarkers of psychiatric treatment response remain elusive. The search for such biomarkers is of particular importance given that subjective ratings of pre-treatment symptom severity often fail to predict treatment outcomes for a range of common psychiatric disorders (e.g., 1). Non-invasive functional magnetic resonance imaging (fMRI) serves as one theoretically viable alternative for prediction of treatment outcomes (e.g., 2). However, typical neuroimaging-based treatment outcome prediction models have been heavily critiqued under the argument that thousands of patients are needed to successfully establish treatment predictors (3). Further, recent meta-analyses demonstrate low overall reliability of both task and resting-state fMRI using standard measures (e.g., functional connectivity and average brain signals) (4, 5). Inaccurate predictions of treatment outcomes will thus necessarily remain (despite large-scale, resource-intensive efforts) if the same unreliable measures continue to be utilized. We need a different approach.

Grossly underappreciated in the clinical domain, evidence continues to mount revealing that moment-to-moment fluctuations in brain activity (i.e., brain signal variability) can viably index the adaptability and effectiveness of neural systems. For example, Garrett and colleagues have shown in a series of experiments that better cognitive performance typically reflects greater moment-to-moment neural variability, the level of which can also be boosted pharmacologically (6–8). Crucially, although the unique predictive power of moment-to-moment neural variability can be more than five times that of conventional mean signal-based approaches (9) and initial evidence for measurement reliability is promising (10), no treatment outcome prediction studies to date have examined brain signal variability.

Here, we provide a first test of the predictive utility of brain signal variability in relation to cognitive-behavioral therapy (CBT) outcomes in social anxiety disorder (SAD) patients. CBT for SAD is an evidence-based treatment intended to limit the avoidance of social situations and reduce self-focused attention - hallmarks of the disorder (11). Although the average group-level effect of CBT can be strong (12), there is considerable variability across-patient response rates, with many SAD patients remaining symptomatic after treatment (13). At its core, CBT is intended to help patients adapt to momentary, social anxiety-provoking demands in the internal and external environment (11). This prompts the appealing question of whether such socially relevant demands may be reflected in moment-to-moment fluctuations in brain signals (i.e., fMRI-variability), and whether brain signal variability could provide a novel predictive signature of CBT treatment outcome in SAD.

To this end, 45 patients with a primary diagnosis of SAD underwent fMRI twice during an 11-week test-retest period before enrollment in a 9-week CBT. We investigated the reliability and predictive power of moment-to-moment neural variability at rest and during a disorder-relevant socio-affective task, while further comparing the predictive accuracy of neural variability to conventional measures of mean neural responses and behavioral self-reports. Our results show that on-task moment-to-moment brain signal variability provides maximal reliability and treatment outcome prediction for SAD.

## Materials and Methods

The study was registered at ClinicalTrials.gov (id: NCT01312571) and ethical approval was obtained from the regional committee at Umeå University, Umeå, Sweden. All participants gave written informed consent prior to participation.

### General procedure

Individuals experiencing social anxiety and seeking treatment were targeted via media advertisements, provided self-reports, and participated in a diagnostic interview as part of the screening. Included participants underwent internet-delivered CBT for SAD for 9 weeks. Before CBT, the patients were subject to a 11-week test-retest period on behavioral self-reports and brain-based fMRI (i.e., baseline 1 and baseline 2). In addition to test-retest reliability, multiple baselines were included to control for standard confounds related to time in study (e.g., regression to the mean and spontaneous remission). Forty-six SAD patients were recruited in the current study. Data from one patient contained outliers, as evidenced by the multivariate mahalanobis distance of brain-behavioral measures (see Supplementary Material Figure S1 for details); thus, our primary results reported here will center on the remaining 45 patients.

### Recruitment of social anxiety disordered patients

Individuals answered online questionnaires on demographics, social anxiety, and depressive and insomnia symptoms as part of the screening. Eligible individuals were interviewed via telephone using (A) the full Mini-International Neuropsychiatric Interview (M.I.N.I.) version 7.0 (14) and (B) the social phobia and major depressive disorder sections of the Structured Clinical Interview for DSM-IV – Axis I Disorders (SCID-I) (15). Included patients were at least 18 years of age, had no neurological disorder, no concurrent psychological treatment, and if treated with a psychotropic medication, they agreed to maintain a stable dose at least 3 months before enrollment and during treatment in the current study. All participants also met magnetic resonance imaging (MRI) safety criteria (e.g., not pregnant, no ferromagnetic objects in the body). At screening, we excluded patients that were currently suffering from ongoing depression (as indexed by scoring >34 on the Montgomery Åsberg Depression Rating Scale, MADRS-S) (16), bipolar or psychotic disorders, alcohol or substance use disorders, or antisocial personality disorder. Further, patients that answered positive to any SAD comorbidity in the M.I.N.I. screening telephone interview were subject to a second face-to-face DSM-5 diagnostic interview. Twenty patients (44.4%, 20/45) had a concurrent psychiatric comorbidity, and four patients (8%, 4/45) were on a stable dosage of selective serotonin reuptake inhibitors (SSRIs), which did not change throughout the study period. Two patients had previously used beta blockers in social situations, but agreed not to use them during the study period. For further study and sample details, see Månsson et al., 2019 (17). See also Table S1 for a detailed summary of demographic and clinical status, and comorbid mental illness. Importantly, all patients that entered treatment also remained throughout the intervention, and took part in post-treatment assessments.

### Cognitive behavioral-therapy (CBT)

Briefly, internet-delivered CBT for SAD is a guided self-help intervention administered over a 9-week period. Each week, the patients are provided with a module containing text and homework assignments based on CBT. The content in this treatment is standardized (i.e., all patients were provided with the same material - identical to our previous RCTs (18, 19)). Patients were in weekly contact with a clinical psychologist, who provided written feedback and guidance via a secured internet platform. The patients undertook a weekly test with questions related to CBT and content of the module. To control for compliance, the patients had to give 100% correct responses on the multiple-choice questionnaire (with the possibility of redoing the test multiple times). After completion of the homework assignments and the multiple-choice quiz, the next module was made available to the patient. Further details regarding internet-delivered CBT have been described extensively elsewhere (20).

Seven clinical psychologists served as therapists in the current study. The mean number (±SD) of years with experience working with CBT was 6.7±5.5 and the allocation of patients to the therapists was randomized. The mean (±SD) number of completed treatment modules was 7.9 (±1.8) and 80% (36/45) of the patients completed at least 7 out of 9 modules.

### Social anxiety, depressive, and insomnia outcomes

The LSAS-SR is a 48-item self-report questionnaire (each question consists of common social situations and the responder is asked to state both his/her anxiety and avoidance in these situations). LSAS-SR is a gold-standard questionnaire to assess treatment-related changes in social anxiety symptoms and the total score typically shows excellent test-retest reliability (e.g., *r* = .83) (21). LSAS-SR was the primary outcome measure of the current study and was administered at multiple times throughout the study period: screening (week 0), first (week 1) and second (week 9) baseline, and immediately after the treatment (week 18). To examine social anxiety as a general construct, secondary social anxiety measures were also collected, including the Social Interaction Anxiety Scale (SIAS) (22), the Social Phobia Scale (SPS) (22), and the Social Phobia Screening Questionnaire (SPSQ) (23). Further, post-treatment interviews were performed via telephone and included SCID-I on SAD (24) and M.I.N.I. on SAD (25), as well as the Clinical Global Impression-Improvement (CGI-I) scale (26). The interviews were performed by two external psychiatrists. Before the interview, the psychiatrists were informed about each patient’s pre-treatment LSAS-SR score, but blind to any post-treatment self-reports. Depressive and insomnia symptoms were assessed using the Montgomery-Åsberg Depression Rating Scale, Self-reported version (MADRS-S) (16), and the Insomnia Severity Index (ISI) (27). Secondary social anxiety outcomes, and depressive and insomnia symptom questionnaires were administered at pre- and post-treatment.

### Neuroimaging

MRI was performed twice for each patient (first and second baseline) and the two sessions were separated by eleven weeks (average number of days between sessions: 77.2±1.6). Also, the time of day each patient was scanned did not vary between the two baselines (average difference in time of day = 1.7±2.3 hours), nor did pre-scan session subjective sleepiness (Karolinska Sleepiness Scale; *B* = 0.13, BSE = 0.25, *Z* = 0.53, *P* = 0.594) (28).

In each scanning session, patients first underwent resting-state and then a task-related condition. Resting-state recordings lasted 340 seconds (sec) and were performed with eyes open (fixation-cross present on screen). As displayed in Figure 1, during the socio-affective face task, patients passively viewed emotional faces (happy/fearful male/female) across two blocks (29). In each block, a single face was repeatedly presented for 200 ms followed by a 300 ms fixation cross for a period of 80 sec (both mini-blocks combined yielded a total of 160 sec of face stimulation). The order of stimuli was counterbalanced across subjects with regard to both the facial expression and sex of the person portrayed in the image. Before (20 sec), in-between (30 sec), and after (20 sec) the face stimulus blocks, fixation blocks were presented.

**Figure 1.**
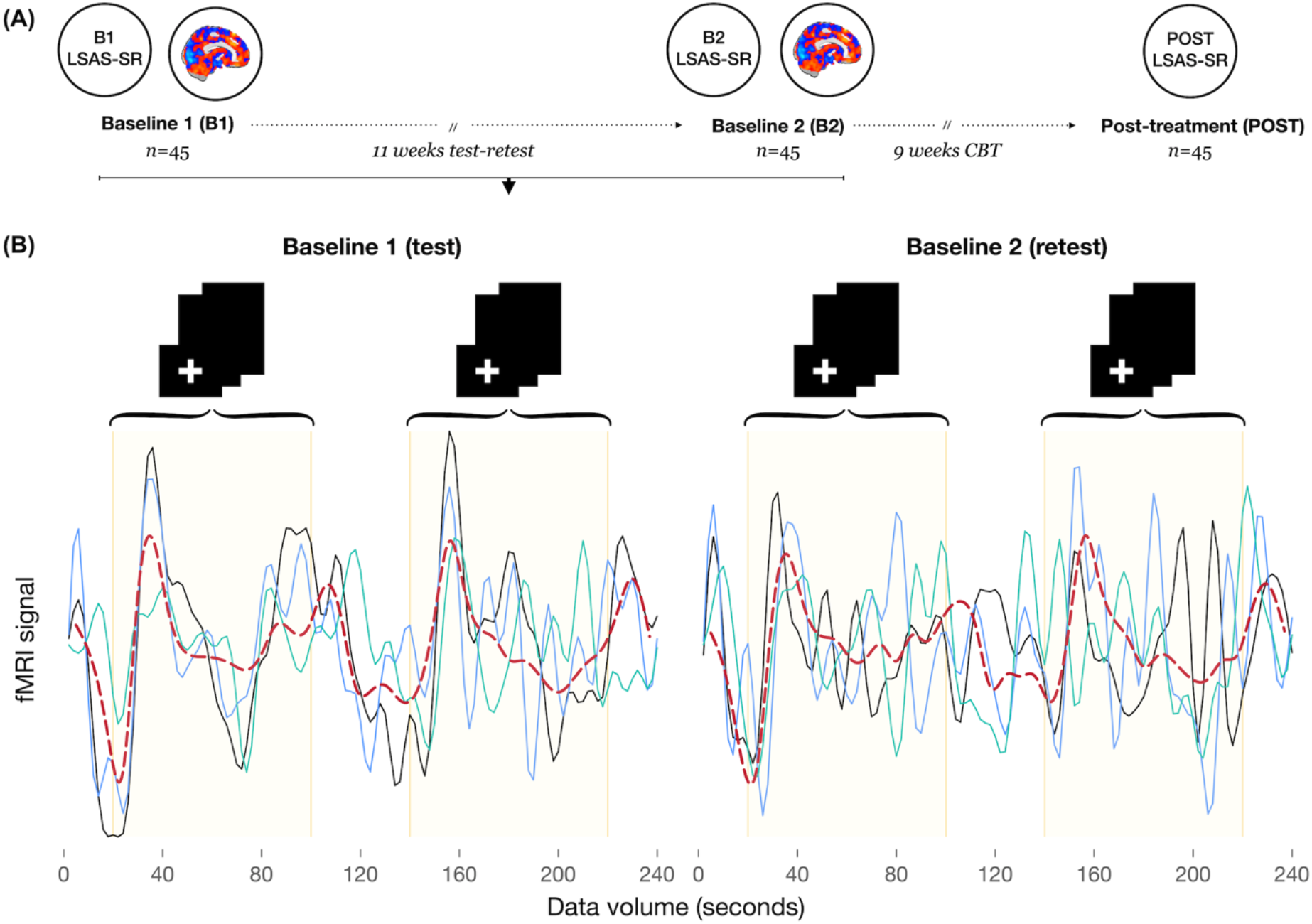
Study design and experimental task. **(A)** Forty-five patients provided behavioral (e.g., LSAS-SR) and brain data (i.e., fMRI) at two time-points: baseline 1 (B1) and baseline 2 (B2) separated by 11 weeks. Further, post-treatment behavioral data after 9 weeks of Internet-delivered CBT were also collected. **(B)** Example of visual cortex (mean-centered) fMRI time series (data volumes in sec) within each baseline session from three random patients (i.e., solid green/blue/black lines). The dashed red line represents the average (median cubic spline) signal across all patients in the current study (*n* = 45). Vertical solid/yellow lines represent stimuli onsets: face 200 ms + 300 ms fixation, with 160 repetitions totalling 80 sec for each block. The non-shaded parts of the time-series represent fixation blocks (i.e., continuous presentation of a fixation cross - data not used). In agreement with the policy of medRxiv, stimuli including faces are not presented in this preprint. **Abbreviations:** CBT, cognitive-behavioral therapy; LSAS-SR, Liebowitz social anxiety scale, self-report version; fMRI, Functional magnetic resonance imaging; Post, Post-treatment;

#### Magnetic resonance imaging

Images were collected on a 3 Tesla General Electrics (GE) scanner with a 32 channel head coil at the Umeå Centre for Functional Brain Imaging (University Hospital of Umeå, Umeå, Sweden). First, an anatomical T1-weighted image (fast spoiled gradient echo) was collected (180 slices, 1 mm thickness, field of view: 250 mm, voxel size: 0.5 × 0.5 × 1 mm^3^) for each patient. Second, for resting-state and task, blood-oxygen-level-dependent (BOLD) contrast images were acquired using the following parameters: 30 ms echo time, 2000 ms repetition time; 80 degree flip-angle, field of view: 250 × 250 mm^3^, matrix size: 96 × 96. Thirty-seven slices with a thickness of 3.4 mm were acquired to capture the whole brain. Ten dummy scans were run before the image acquisition started to avoid signals resulting from progressive saturation.

Stimuli were presented on a computer screen and seen by the participant through a mirror attached to the head coil. Headphones and earplugs were used to reduce perception of scanner noise and cushions in the head coil reduced movement. Experienced MRI nurses were taking care of all participants.

#### Brain image preprocessing pipeline

fMRI data were preprocessed with FSL 5 (30, 31). Pre-processing included motion-correction, initial bandpass filtering (.01–.10 Hz), and detrending (up to a cubic trend) using SPM12. We also utilized extended preprocessing steps to further reduce potential data artifacts (6, 8, 9). Specifically, we subsequently examined all functional volumes for artifacts via independent component analysis (ICA) within-run, within-person, as implemented in FSL/MELODIC (32). Noise components were identified according to several key criteria: A) Spiking (components dominated by abrupt time series spikes); B) Motion (prominent edge or “ringing” effects, sometimes [but not always] accompanied by large time series spikes); C) Susceptibility and flow artifacts (prominent air-tissue boundary or sinus activation; typically represents cardio/respiratory effects); D) White matter (WM) and ventricle activation (33); E) Low-frequency signal drift (34); F) High power in high-frequency ranges unlikely to represent neural activity (≥ 75% of total spectral power present above .10 Hz;); and G) Spatial distribution (“spotty” or “speckled” spatial pattern that appears scattered randomly across ≥ 25% of the brain, with few if any clusters (i.e., ∼20 voxels at 3 × 3 × 3 mm voxel size).

Examples of these various components we typically deem to be noise can be found in previous work (10). By default, we utilized a conservative set of rejection criteria; if manual classification decisions were challenging due to mixing of “signal” and “noise” in a single component, we generally elected to keep such components. ICA components were reviewed by an experienced MRI research engineer and ambiguous noise/signal ICs were discussed within the research group to reach common decisions. Components identified as artifacts were then regressed from corresponding fMRI runs using the regfilt command in FSL. Finally, we registered functional images to participant-specific T1 images, and from T1 to 3 mm standard space (MNI 152) using FLIRT (affine). Finally, we masked the functional data with the GM tissue prior provided in FSL (probability > 0.37).

#### Voxel-wise estimation of brain signal variability

For resting state, SD_BOLD_ was computed across the entire denoised time series for each voxel. To calculate SD_BOLD_ for the socio-affective face task, we also performed a block normalization procedure to account for residual low frequency artifacts (as in previous work) (6). We first normalized all blocks for the socio-affective face task such that the overall 4D mean across voxels and blocks was 100, within-person. For each voxel, we then subtracted the block mean, concatenated across all task blocks, and computed voxel SD_BOLD_ across this concatenated time series (35).

We also sought to compare SD_BOLD_ results to a more typical mean-based measure of fMRI activity (MEAN_BOLD_) during the socio-affective face task. Accordingly, we calculated MEAN_BOLD_ by first expressing each within-block volume as percent change from the average of the ten preceding (fixation) block scans, calculating mean percent change within each block, and averaging across all face blocks (a typical method in the partial least squares (PLS) data-analysis framework; see below for model details and implementation).

### Statistical modeling

#### Treatment outcomes and predictors of clinical outcome

Longitudinal behavioral data were implemented in repeated measure analyses with generalized estimating equations (default gaussian and exchangeable correlation structure) to calculate clinical outcomes across time (i.e., screening, first and second baseline, and post-treatment). Within-group Cohen’s *d* effect sizes were calculated by dividing the mean difference with the standard deviation and correcting for the correlation between time-points (i.e., post-treatment vs screening). Simple and hierarchical linear regressions were used to regress predictor (s) on clinical outcomes. Pre-treatment LSAS-SR as a predictor was compared with all brain-derived variables. Comparisons between predictive brain models were realized using multiple regression models and the adjusted *R*^2^ values are reported throughout the paper. All correlations were performed in a parametric way (Pearson correlations) and nonparametric bootstrapping (× 1000) was used to estimate standard errors (BSE) and 95% confidence intervals of regression models. Further, permutation tested statistics (× 1000) are reported. STATA Statistical Software (v15.1, STATA Corporation, College Station, TX, USA) was used.

#### Estimating brain-based correlates of clinical outcome

To examine the relationship between BOLD activity and treatment success, we utilized a multivariate PLS analysis (36, 37) (implemented in MATLAB 9.6.0.1072779; R2019a; Natick, Massachusetts: The MathWorks Inc. 2018). In brief, our PLS model began with a correlation matrix that captured the between-subject correlation (Pearson’s *r*) of brain activity (e.g., SD_BOLD_) in each voxel (51 609 per subject) and subject-wise delta LSAS-SR score (delta Post-B1). Next, this correlation matrix was decomposed using singular value decomposition (SVD), which yielded brain-based saliences (*k* = 51 609) that represent the topographically resolved correlation strength between BOLD activity (e.g., SD_BOLD_) and delta LSAS-SR scores. For every subject, so-called “brain scores” were then calculated through the dot product of these weights with voxel-wise BOLD values. To estimate the robustness of model weights (saliencies), 1000 bootstraps with replacement were utilized, and the division of each voxel-wise salience by its corresponding bootstrap standard errors yielded pseudo-z normalized estimates called bootstrap ratios (BSRs). The topographical pattern of BSRs reveals how the correlation between brain activity (e.g., SD_BOLD_) and behavior (delta LSAS-SR) is distributed throughout the brain. Furthermore, bootstraps were used to estimate 95% confidence intervals (CIs) for observed correlations between brain measures and delta LSAS-SR scores.

To compare relative predictive utility and reliability, this PLS approach was utilized separately to test the link between a series of different brain measures (i.e., socio-affective face task-based SD_BOLD_, face task-based MEAN_BOLD_, and resting-state SD_BOLD_) and treatment-related LSAS-SR changes. We then ran a series of PLS models to also examine the influence of data volume on the strength and reliability of brain-based effects (i.e., for task-based SD_BOLD_ and MEAN_BOLD_: first 40, first 80, and all 160 sec; for resting-state SD_BOLD_: first 40, first 80, first 160, and all 340 sec). The same behavioral variable (the difference in LSAS-SR scores between the initial measurement (Baseline 1) and post-treatment (Post-B1)) was used in all of these models.

##### Cross-validation framework for brain and behavioral predictors of treatment outcome

One key goal in the present work is to establish and compare the relative strength and reliability of different brain and behavioral predictors of treatment outcome. To do so, we employed a custom two-step reliability-based cross-validation framework (see Figure 2).

**Figure 2.**
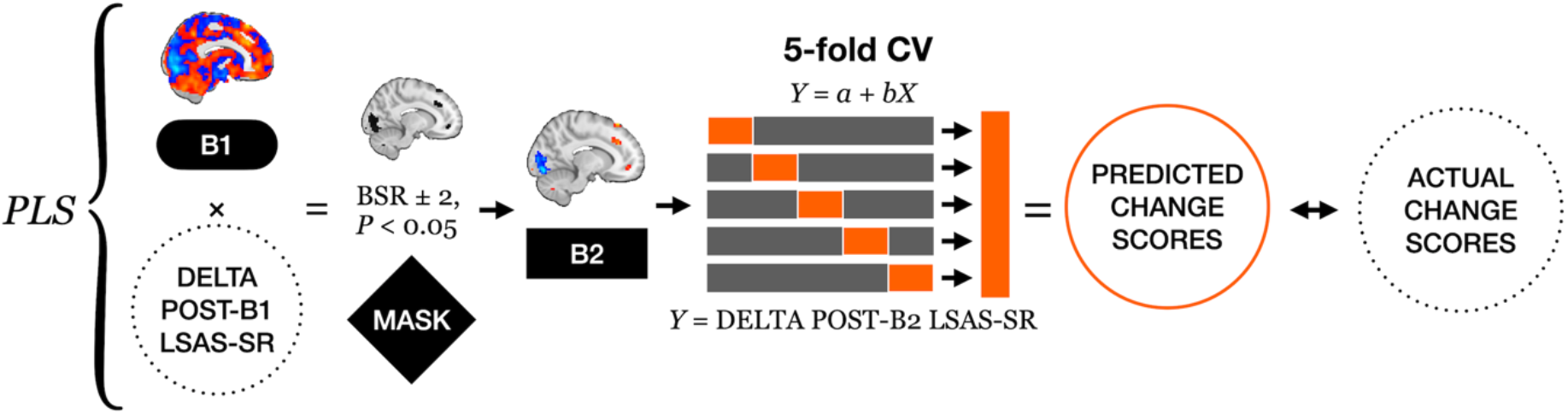
A general overview of the analytic framework. The analysis of neuro-behavioral correlations between BOLD signals at B1 and the treatment outcome (i.e., delta LSAS-R score post-treatment minus B1) was computed for all 45 subjects via behavioral PLS. A bootstrap-based mask was created (BSR ± 2) based on B1 data. To compute “reliability-based cross validation”, weights from this mask were applied to corresponding voxels in B2 brain data to permit extraction of subject-specific brain scores at B2 (i.e., no additional PLS model was run on B2 data). Prediction of treatment outcome-based change scores was then performed using 5-fold cross-validation, from which empirical treatment-based change scores were correlated with predicted scores. **Abbreviations:** LSAS-SR, Liebowitz social anxiety scale, self-report version; fMRI, Functional magnetic resonance imaging; CBT, Cognitive behavior therapy; CV, Cross-validation; BSR, Bootstrapped ratio; PLS, Partial least squares; B1, Baseline 1; B2, Baseline 2; Post, Post-treatment;

First, as described above (separately for each brain measure and data volume), we computed PLS models linking brain activity and reductions in LSAS-SR scores of all participants based on the first baseline (delta Post-B1) MRI recording. Second, the resulting voxel-wise BSRs for each model were thresholded at ±2 while excluding all clusters smaller than 20 voxels. Third, we applied these weights to MR data recorded during the second baseline measurement (B2; 11 weeks after B1) to extract subject-specific brain scores without re-estimating PLS models. Doing so permits a form of “metric invariance” (38) (here, the fixing of model weights between the two baseline periods), allowing for a clearer and more direct comparison of both measurement points. Finally, these B2 brain scores were used to estimate linear relationships between BOLD activity and changes in LSAS-SR scores (delta Post-B2) within a 5-fold cross validation framework. Linear coefficients were estimated on a subset of subjects (training set, *n* = 36) before being applied to another, non-overlapping subset of subjects (test set, *n* = 9), based on which predictions were generated.

While our reliability-based modeling approach does not, by definition, seek out-of-sample prediction *per se* due to the common topographical weight map estimated at B1 and fixed at B2 (see above), we nevertheless test for “generalizability” of our models on two intertwined levels. First, because PLS weights from the B1 MR measurement are applied to B2 MR data without re-running PLS, any statistically meaningful prediction of treatment success can *only* emerge if the link between BOLD activity and treatment outcome is similar in both magnitude and topographical distribution across two completely separate MR recordings 11 weeks apart. Second, our 5-fold cross validation approach further limits model overfitting (in the presence of our limited sample size). By directly comparing the strength and reliability of different brain measures for treatment prediction here, future work on larger-scale out of sample prediction of treatment outcomes can then proceed.

##### Prediction accuracy estimation

To compare the predictive power of pre-treatment LSAS-SR and BOLD fMRI-derived (task SD_BOLD_, task MEAN_BOLD_, and resting-state SD_BOLD_) variables, we calculated the Pearson correlation between predicted and observed LSAS-SR changes. Additionally, 95% confidence intervals were estimated for all correlation coefficients using a bootstrap approach (2500 bootstraps). Furthermore, and to offer a second metric for the relative comparison of predictors, we calculated the mean absolute scaled error (MASE) (39), which is defined as the ratio of the mean absolute prediction error to the mean absolute error of the one-step naïve forecast. While a MASE value of 1 represents equal predictive power of naive forecast and another predictor of interest, values below 1 depict a predominance of the predictor of interest where the improvement in prediction accuracy is 1–MASE %. MASE offers a scale-invariant measure of prediction accuracy and hence is directly comparable across different predictors regardless of their scale. Importantly, MASE penalizes over- and under-forecasting (i.e., too high vs. too low predicted scores, respectively) equally, rendering it a symmetric measure of prediction error. As other commonly used metrics of prediction accuracy do not offer scale independence or symmetry (e.g., root mean squared error, RMSE), MASE has been suggested as an ideal measure to compare the accuracy of different predictions (40).

#### Standard test-retest reliability estimation

Intraclass Correlation Coefficients ICC (C,1) based on the degree of consistency among measurements were calculated to determine test-retest reliability on self-reported behavioral and brain-based variables (41) between the two baseline measurements (separated by 11 weeks). MATLAB 9.7.0.1190202 (R2019b; Natick, Massachusetts: The MathWorks Inc.; 2018) was used to compute ICCs (42). Between B1 and B2 measurements, ICCs were calculated on LSAS-SR (total score), and for each brain measure and data volume, on PLS brain scores and on all voxels across the whole-brain (*k* = 51 609). Bootstrapped (× 1000 bootstraps) lower and upper bound 95% confidence intervals are reported. The ICCs were categorized as poor < 0.40, fair = 0.40 to 0.59, good = 0.60 to 0.74, or excellent ≥ 0.75 according to Cicchetti and Sparrow (43).

#### Data and code availability

All code and statistical software commands will be available at https://github.com/LNDG. Due to current ethics constraints, we cannot at present make the raw data openly available, but please contact the first author (K.M.) to discuss other potential routes to data access.

## Results

### Treatment outcomes

The primary social anxiety outcome (LSAS-SR) decreased 33.46 points on average from screening to post-treatment (see Figure 3A). Large within-group Cohen’s d effect sizes (>1.49) were observed for both LSAS-SR and secondary outcomes (i.e., SIAS, SPS, SPSQ), all permuted *P*s <0.001. The clinician-administered CGI-I interviews found 32 of 45 patients’ mental health to be much (48.9%) or very much (22.2%) improved at post-treatment. Despite the overall improvement, only a minority of patients were in probable remission at post-treatment, as indicated by a score less than 30 on LSAS-SR (28.9%, 13/45) or by being free from SAD according to DSM-5 criteria (17.8%, 8/45). See Tables S2-S3 and Figures S2-S4 for a detailed presentation of all clinical outcomes.

**Figure 3.**
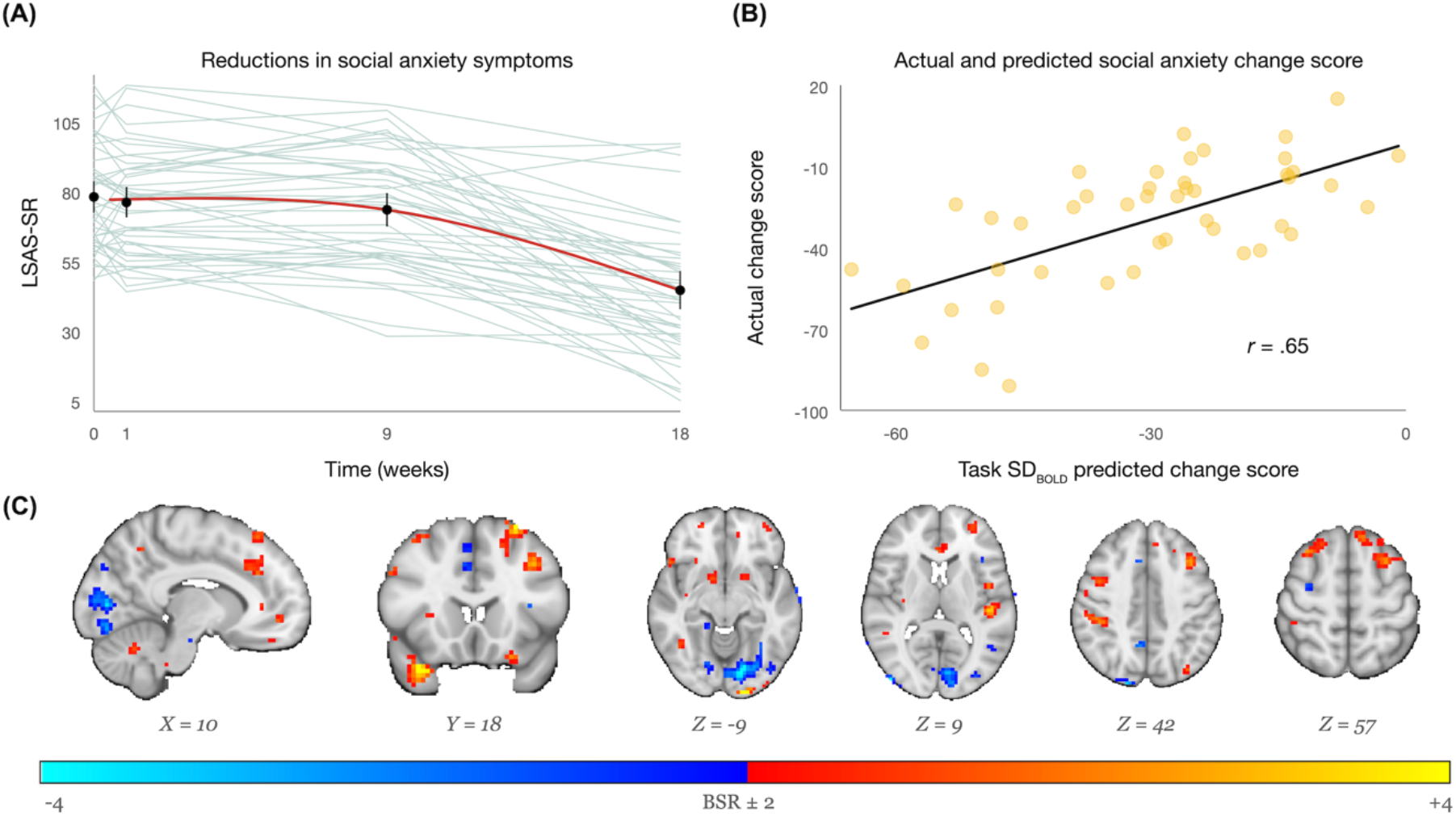
Reductions in social anxiety symptoms and task-related brain signal variability as a predictor of treatment outcome. **(A)** Change in the primary social anxiety outcome LSAS-SR from screening, baseline 1, baseline 2 to post-treatment. The solid line represents the median cubic spline. **(B)** Task-based SD_BOLD_ predicted treatment change score is strongly related to empirical change scores. **(C)** Task-based SD_BOLD_ spatial pattern reflecting treatment outcome. Blue regions = lower SD_BOLD_ associated with better treatment outcome; yellow/red regions: higher SD_BOLD_ associated with better treatment outcome. *X Y Z* below the brains represent MNI coordinates. Further, an unthresholded statistical maps are available at NeuroVault.org (https://identifiers.org/neurovault.collection:9030). **Abbreviations:** LSAS-SR, Liebowitz social anxiety scale, self-report version; SD_BOLD_, standard deviation of BOLD; BOLD, Blood-oxygen-level-dependent imaging;

### Task-related brain signal variability strongly predicts treatment outcome

Moment-to-moment brain signal variability during emotional face processing robustly predicted social anxiety change scores (post-pre CBT) (5-fold cross-validated; *r (45)*_ACTUAL,PREDICTED_ = .65, MASE = .54, permuted *P* < 0.001; Figure 3B). Specifically, low signal variability in the right visual cortex, and high variability in the anterior cingulate, medial prefrontal, and temporal cortices predicted larger reductions in social anxiety symptoms (see Figure 3C, Figures S5-S8 and Tables S4-S8 for a complete presentation of neural activations and data density plots). The predictive power of task-related SD_BOLD_ remained nearly identical even when the data volume was reduced by either 50% (80 sec; *r (45)*_ACT,PRED_= .65, MASE = .53, permuted *P* < 0.001) or 75% (40 sec; *r (45)*_ACT,PRED_ = .62, MASE = .52, permuted *P* < 0.001) - see also Figure 4 and Tables S9-S10 for details of various multiple regression models spanning brain measures and data volumes.

**Figure 4:**
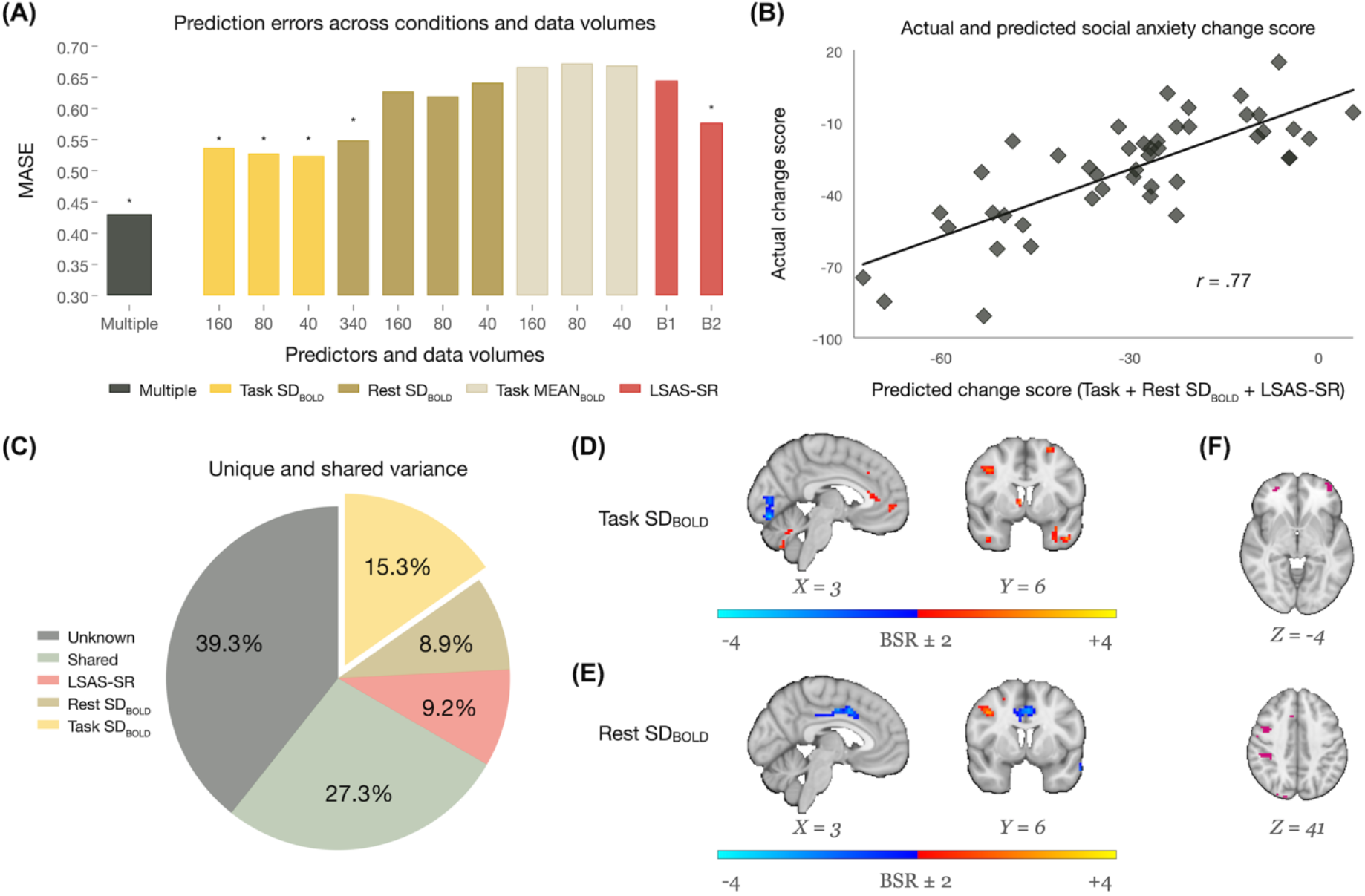
Treatment outcome prediction accuracies and brain signal variability. **(A)** Prediction accuracy (i.e., MASE) for each condition (i.e., task SD_BOLD_, resting-state SD_BOLD_, task MEAN_BOLD_, LSAS-SR at B1 and B2) and across data volumes (i.e., 40, 80, 160 and 340 sec). * denotes significant (permuted *P* < 0.001) zero-order prediction models noted in Table S4. Lower values indicate better model performance. **(B)** 5-fold cross-validated correlation between empirical and predicted treatment change scores using all zero-order significant conditions (i.e., 160 sec task SD_BOLD_, 340 sec resting-state SD_BOLD_, and LSAS-SR at B2). **(C)** Unique and shared variance (*R*^2^) between model predictors and treatment outcome. **(D)** Task-related SD_BOLD_ and **(E)** resting-state SD_BOLD_ spatial pattern reflecting treatment outcome. Blue regions represent less variability predicting better outcome, whereas yellow/red regions represent higher fMRI signal variability predicting better outcome. **(F)** Displays overlapping activations between task SD_BOLD_ (160 sec) and resting-state SD_BOLD_ (340 sec). In figure F, the spatial pattern for each condition represents a binary mask. *X Y Z* below the brains represent MNI coordinates. Unthresholded statistical maps will be available at NeuroVault.org (https://identifiers.org/neurovault.collection:9030). **Abbreviations:** MASE, Mean absolute scaled error; LSAS-SR, Liebowitz social anxiety scale, self-report version; B1, Baseline 1; B2, Baseline 2; SD_BOLD_, Standard deviation of BOLD; MEAN_BOLD_, Average BOLD activity; BSR, Bootstrap ratio; BOLD, Blood-oxygen-level-dependent imaging;

### Comparing the predictive utility of task-based fMRI variability to resting-state variability, mean activity, and self-reports

In a multiple regression model including all potential behavioral and brain-based predicted social anxiety change scores (and equal data volumes of 160 sec for all brain measures), task-related SD_BOLD_ (*β* =.61, permuted *P* < 0.001) dominantly outperformed (A) resting-state SD_BOLD_ (*β* = .26, permuted *P* = 0.090), (B) task-related MEAN_BOLD_ (*β* = -.07, permuted *P* = 0.621), and (C) pre-treatment social anxiety severity (second baseline LSAS-SR, *β* = .22, permuted *P* = 0.186). The model accounted for 54% of the variance in the social anxiety change score. Although self-reported social anxiety at pre-treatment did not predict outcome within our full model, a moderate zero-order correlation did indicate that more severe patients exhibited greater reduction in social anxiety (*r (45)*_ACT,PRED_ = .45, permuted *P* < 0.001) - see also Table 1 and Figure 4A for a complete presentation of statistical results. In a second model including the same predictors, but instead using the brain score from the full available resting-state SD_BOLD_ data volume (340 sec). Here, we found that resting-state SD_BOLD_ also uniquely predicted treatment outcome (*β* = .34, permuted *P* = 0.039), but task-related SD_BOLD_ remained the strongest treatment outcome predictor (*β* = .41, permuted *P* = 0.018) despite less than half the data volume (160 sec) of resting-state. Furthermore, as displayed in Figure S9, we demonstrate equal model performance without bootstrap-based thresholding based on the baseline 1 data (i.e., no feature selection). All peak neural activations (i.e., task SD_BOLD_, resting-state SD_BOLD_, task MEAN_BOLD_) and corresponding coordinates are reported in Tables S6-S8, and unthresholded statistical maps for task SD_BOLD_ and resting-state SD_BOLD_ (the only significant brain-based predictors) are available at NeuroVault.org (https://identifiers.org/neurovault.collection:9030).

**Table 1.** Univariate (zero-order) and multiple predictor models of treatment outcome across different within-patient data volumes. See Supplementary Material Table S4 for a complete presentation of model results across all data volumes.

| Pre-treatment predictors | Data volume | MASE | $r$ (45) | 95% CI | | Permuted $P$ |
| --- | --- | --- | --- | --- | --- | --- |
|  |  |  |  | Lower | Higher |  |
| Behavioral |  |  |  |  |  |  |
| LSAS-SR at B1 | 48 items | 0.65 | .27 | -.01 | .56 | 0.071 |
| LSAS-SR at B2 | 48 items | 0.58 | .45 | .20 | .70 | <0.001* |
| Brain |  |  |  |  |  |  |
| Task SD<sub>BOLD</sub> | 160 sec | 0.54 | .65 | .51 | .79 | <0.001* |
| Rest SD<sub>BOLD</sub> | 160 sec | 0.63 | .19 | -.08 | .46 | 0.204 |
|  | 340 sec | 0.55 | .55 | .35 | .75 | <0.001* |
| Task MEAN<sub>BOLD</sub> | 160 sec | 0.67 | -.18 | -.49 | .12 | 0.218 |
| Brain SD<sub>BOLD</sub> and behavioral self-reports combined |  |  |  |  |  |  |
| Multiple predictors** |  | 0.43 | .77 | .66 | .89 | <0.001 |

\*Significant zero-order prediction. \*\*The multiple regression model includes all significant zero-order predictors (i.e., 160 sec task $SD_{BOLD}$ , 340 sec resting-state $SD_{BOLD}$ , and LSAS-SR at B2). **Abbreviations:** MASE, Mean absolute scaled error; LSAS-SR, Liebowitz social anxiety scale, self-report version; B1, Baseline 1; B2, Baseline 2; $SD_{BOLD}$ , neural variability; $MEAN_{BOLD}$ , average neural response; Sec, Seconds; BOLD, Blood-oxygen-level-dependent imaging;

As displayed in Figure 4A, a final cross-validated model including only the univariate significant predictors of treatment outcome (i.e., task SD_BOLD_ (160 sec), resting-state SD_BOLD_ (340 sec), and second baseline LSAS-SR), improved the predictive accuracy beyond any single predictor (*r (45)*_ACT,PRED_ = .77, MASE = .43, permuted *P* < 0.001; see also Table 1). However, the unique variance associated with task-based SD_BOLD_ was notably higher than all other predictors (Figure 4C). Furthermore, the spatial patterns capturing neuro-behavioral correlations of task-based and resting-state SD_BOLD_ differed considerably (see also Figure 4F, and Figure S10), suggesting that the contribution of each SD_BOLD_ estimate to treatment outcome prediction is complementary both statistically (with regard to effect size and MASE values) and with regard to involved brain regions.

Neither depression nor insomnia severity at pre-treatment predicted social anxiety treatment outcomes (all permuted *P*s > 0.188). Similarly, treatment credibility ratings, demographics (as displayed in Table S1: age, sex, education, marital status), or psychiatric comorbidity (yes/no) did not predict treatment outcome (all permuted *P*s > 0.071).

### Task-based signal variability prediction specifically generalizes across secondary social anxiety measures

As the task SD_BOLD_ condition was the strongest treatment outcome predictor, we investigated if this predictor also generalized across secondary social anxiety outcomes. To do so, we first estimated a single PCA component representing all available secondary outcomes (i.e., self-reported SIAS, SPS, SPSQ, and the clinician-administered CGI-I; eigenvalue = 2.80, 70% explained variance and component loadings varied from .72 to .89). This component score correlated strongly with the task-based SD_BOLD_ predicted social anxiety change score used in our primary (LSAS-SR) analyses above (*R*^2^ = 34%, permuted *P* < 0.001). Further, as Table S2 and Figure S3B depict, symptoms of depression (permuted *P <* 0.001) and insomnia (permuted *P* = 0.042) decreased over the course of therapy. However, the socio-affective face task SD_BOLD_ predicted social anxiety change score was not associated with reductions in depressive or insomnia symptoms (all permuted *P*s > 0.351). Taken together, these results support both sensitivity and specificity of our task-based SD_BOLD_ prediction model of treatment outcome.

### Eleven-week test-retest reliability

As displayed in Figure 5 and Table S11, the eleven-week test-retest reliability (Baseline 1 *versus* Baseline 2) was excellent both for the primary social anxiety measure (LSAS-SR; ICC_B1,B2_ = 0.84, CI 95% 0.75, 0.90) and our task-related SD_BOLD_ measure (PLS brain scores; ICC_B1,B2_ = 0.80, CI 95% 0.68, 0.87). Task SD_BOLD_ ICC values dropped but remained reasonable after reducing data volumes to 80 sec (ICC_B1,B2_ = 0.76) and 40 sec (ICC_B1,B2_ = 0.62). In contrast, task-related MEAN_BOLD_ showed very poor reliability (all ICC’s_B1,B2_ ∼0), indicating that the MEAN_BOLD_ prediction model was not useful here. When data volume was equated across conditions, resting-state SD_BOLD_ also showed less reliability than task-related variability, and only when the full resting-state data volume was examined (340 sec, representing more than double the available task SD_BOLD_ data volume) did reliability improve to the level achieved by the 160 sec task SD_BOLD_ (resting-state ICC_B1,B2_ = 0.81, CI 95% 0.74, 0.88). Voxel-wise ICC’s are displayed and reported in Table S12.

**Figure 5:**
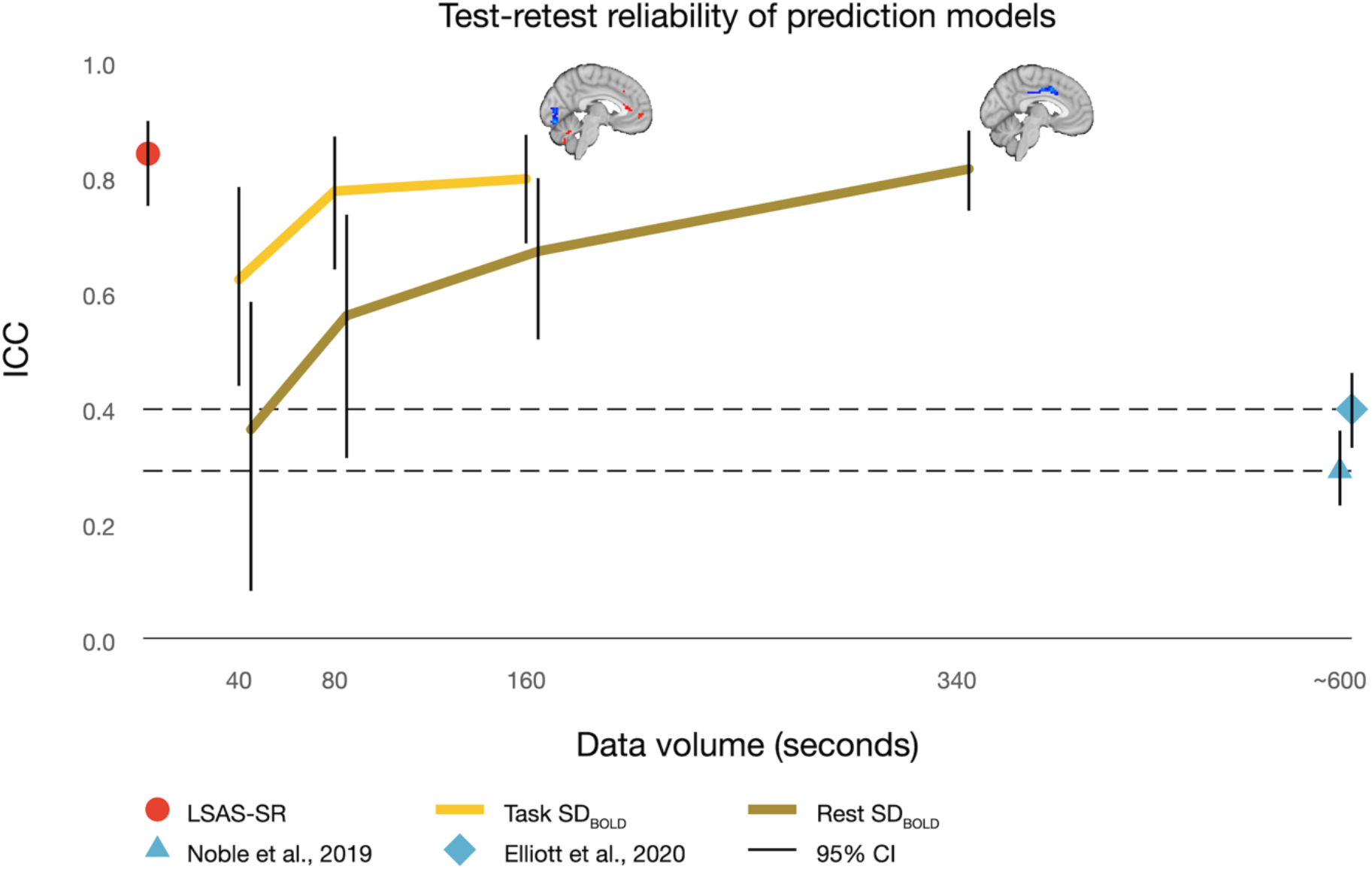
Test-retest reliability. ICCs were computed for LSAS-SR scores as well as fMRI BOLD across conditions (i.e., task SD_BOLD_ and resting-state SD_BOLD_) and data volumes (i.e., 40, 80, 160 and 340 sec). Error bars represent bootstrapped, bias-corrected 95% confidence intervals. For reference, two meta-analyses on test-retest reliability using conventional analytics (i.e., functional connectivity and average brain signals) are presented. Noble et al., 2019 (ref 4) and Elliott et al., 2020 (ref 5) are meta-analyses on standard measures of task- and resting-state fMRI. **Abbreviations:** ICC, Intraclass correlation coefficient; SDBOLD, neural variability; BOLD, blood-oxygen-level-dependent imaging; fMRI, functional magnetic resonance imaging; LSAS-SR, Liebowitz Social Anxiety Scale. Self-report version;

## Discussion

In the present study, we found that internet-delivered CBT successfully reduced SAD patients suffering (LSAS-SR_PRE-POST_ Cohen’s *d* = 1.62) and that pre-treatment brain signal variability was an accurate and reliable predictor of treatment outcome. A multiple predictor model that included task-based SD_BOLD_, resting-state SD_BOLD_, and pre-treatment social anxiety severity showed excellent prediction accuracy (*R*^2^ = 61%, MASE = .43). Task-related SD_BOLD_ was the strongest predictor and exhibited excellent reliability. This relatively short (160 sec) estimate of task-based BOLD signal variability outperformed resting-state SD_BOLD_, standard MEAN_BOLD_, and pre-treatment self-reported social anxiety. Resting-state variability also uniquely predicted treatment outcome in our full model, but accounted for ∼50% less unique explained variance than task-based SD_BOLD_ and required more than double the data volume to achieve comparable reliability and treatment outcome prediction accuracy. Crucially, socio-affective face task SD_BOLD_ was both sensitive and specific to social anxiety treatment outcomes.

### Estimating moment-to-moment fluctuations during simple, disorder-relevant tasks may help optimize treatment prediction

Clinical neuroscientists often argue that resting-state neuroimaging protocols are preferable for ease of implementation and minimization of demands on patients. Our robust task-based treatment prediction results were achieved using a disorder-relevant task (socio-affective visual processing in SAD patients) with extremely low cognitive requirements (passive viewing, no behavioral responses required) and absolutely minimal scan time (2 minutes 40 sec, far shorter than typical resting-state scans). Further, the spatial patterns for task and resting-state SD_BOLD_ were largely distinct. Thus, while both task and resting-state variability contributed to CBT outcome prediction, the two measurements represent different neural signatures. If simple, demand-minimal fMRI remains a primary goal for biomarker development in psychiatry, passive, disorder-relevant tasks should be included in future large-scale studies of treatment outcome, particularly when BOLD fluctuations can be examined. Here, we employed a simple and straightforward calculation of each patient’s brain signal variability, for which code is freely available and deployable for use in the majority of already collected patient fMRI data.

### Moving beyond average neural signals for reliable treatment prediction

Our task-based SD_BOLD_ prediction model also dominated a more conventional analytic approach using mean brain signals (MEAN_BOLD_; i.e., the average fMRI signal across time) to estimate treatment outcomes. Why might MEAN_BOLD_ perform so poorly? Recently, alarming meta-analyses demonstrate that the average ICC may be as low as .40 (95% CI = .33, .46) for common experiments using conventional mean-based analyses in task-based fMRI (5). Crucially, the test-retest reliability of such standard (and alternative) fMRI measures in the treatment outcome prediction literature remains largely unknown. Our task-based SD_BOLD_ treatment prediction model demonstrated excellent 11-week test-retest reliability (ICC_B1,B2_ = .80), and even with minimal data (40 sec), task SD_BOLD_ was far more reliable (ICC = .62) than MEAN_BOLD_ when all available data (160 sec) were used. Beyond the poor performance of MEAN_BOLD_ here, another meta-analysis also revealed very low reliability (ICC = .29; 95% CI = .23, .36) for connectivity-based analyses of resting-state fMRI data (4). Utilizing brain measures with such poor reliability may continue to contribute to non-replicability, and we argue that moment-to-moment brain signal variability computations are therefore strong candidates for future smaller- and larger-scale investigations of biomarkers in psychiatric research and treatment outcome prediction.

### What could BOLD variability reveal about social anxiety treatment outcomes?

Researchers often conceive signal variability as unreliable and unwanted “noise.” However, moment-to-moment brain variability continues to exhibit a host of behaviorally- and group-relevant effects in cognitive neuroscience (e.g., for a review see (44)), yet remains grossly underutilized in clinical research. To our knowledge, SAD has not previously been linked to BOLD variability. It has been argued that an individual’s brain signal variability may (A) reflect available neural dynamic range for the veridical/accurate processing of incoming stimuli (45) and (B) index a more cognitively effective system overall (44). One characteristic SAD symptom is self-focused attention; as a result of an external socio-affective “trigger”, SAD patients become self-attentive and biased towards internal cognitive and emotional processes, leading to deficits in the ability to disengage from internally-focused modes (11). As such, SAD patients may indeed “filter” external socio-affective stimuli through their own internal biases, showing a heightened focus on socio-affective content at the cost of an incomplete representation of such stimuli. CBT includes cognitive and behavioral interventions for dealing with excessive anxiety, such as shifting one’s attention from internally referenced processing toward a more faithful representation of external input. Previous work has shown that lower signal variability in the visual cortex should be expected when individuals *do not* fully process the complexity of visual input (45). It is plausible that treatment-responsive SAD patients express more limited visuo-cortical brain signal variability due to a relative inability to fully process external socio-affective stimuli, a function that may be directly improved via CBT. Complementarily, we also found that patients who displayed *higher* variability in the prefrontal cortex profited more from CBT. Past work consistently shows that greater BOLD variability in the frontal lobes typifies healthy, higher performing adults across a host of different cognitive domains, such as attentional capacity, working memory, and verbal abilities (6, 8–10, 46, 47) (for a review see (44)). Accordingly, prefrontal signal variability may be required to respond to internet-delivered CBT, a treatment process that requires self-motivated learning, working memory, and verbal capacity. Although these interpretations remain speculative, novel questions related to intra-individual variability could be key for future directions in neuroscience-based psychiatric research. To best do so, future longitudinal studies are needed that mechanistically investigate the links between joint changes in neural variability and psychiatric treatment outcomes.

### Limitations and future directions

We provide first evidence for within-sample reliability-based mapping of a series of different fMRI-based measures and experimental conditions in relation to treatment outcome. However, ultimately, the utility of any prediction model is determined by its ability to generalize to new unseen patients. Poldrack and colleagues (3) recently criticised current prediction practices in the neuroimaging literature (e.g., 48– 51), claiming that hundreds or even thousands of patients are needed for out of sample prediction. Even when such high data volume is available, prediction using conventional brain measures can work (*N* = 1 188) (2), but replication is not guaranteed in completely independent patient samples (52). We provide clear, reliability-based evidence for the importance of considering measures of brain signal variability in treatment prediction, and we argue that the use of such reliable tools may markedly reduce the need for such massive, resource-intensive samples for treatment prediction. Further, although accurate and reliable prediction of response to single treatments is a first crucial step for the field, it is essential to know the *type* of treatment to which a particular patient is more likely to respond to achieve maximum clinical utility. To achieve this, a reliable and accurate treatment outcome predictor is needed, and herein we present fMRI variability as a strong candidate for future investigations. Finally, our socio-affective face task SD_BOLD_ treatment outcome prediction model was specific to social anxiety. However, there is well-established evidence that moment-to-moment brain signal variability is linked to various state- and trait-related functions in different samples (8, 44) and it is thus possible that BOLD variability may also be sensitive to a variety of different treatment outcomes in other common psychiatric disorders.

### Summary

In conclusion, neural variability has the potential to offer unique insights into factors that affect patient responses to psychiatric treatments. Here, we demonstrate that intra-individual variability in neural response is a reliable and accurate predictive biomarker of treatment success, even when using a simple passive task administered in under 3 minutes. Ultimately, our findings may help improve precision medicine and clinical decision-making in psychiatric populations.

## Data Availability

All code and statistical software commands will be available online (https://github.com/LNDG/). Due to ethics constraints, we cannot at present make the raw patient data openly available; please contact the first author (K.M.) to discuss potential routes to data access.

## Acknowledgements

This study was performed at the Centre for Functional Brain Imaging (UFBI), Umeå University and the University Hospital of Umeå, and we kindly thank Rebeca de Peredo Axelsson, Hans-Olov Karlsson, Kerstin Englund, Mikael Stiernstedt, Lars Nyberg and Carl-Johan Boraxbekk for support. Gerhard Andersson, Martin Kraepelien, Lise Bergman Nordgren, Erik Hedman-Lagerlöf, Samir El Alaoui and Jens Högström provided state-of-the-art clinical expertise. Cecilia Svanborg and Josef Isung contributed with clinical interviews. The MRI data collection was supported by Nils Hentati Isacsson and Örn Kolbeinsson - you are the best! Thanks also to the Max Planck Institute for Human Development, Lifespan Neural Dynamic Group team members for support in adapting and reviewing code: Steffen Wiegert, Marija Tochadse, Alexander Skowron. Last but not least, a warm thanks to all participants in this study!

We kindly thank the generous research grants K.M. and T.F. received from the Swedish Research Council (2018-06729, and 2016-02228), and the Swedish Brain Foundation (FO-2016-0106). D.G. and K.M. were supported partially by an Emmy Noether Programme grant from the German Research Foundation to D.G. and by the Max Planck UCL Centre for Computational Psychiatry and Ageing Research in Berlin. The funders had no role in study design, data collection and analysis, decision to publish, or preparation of the manuscript.

## Author Contributions

K.M., T.F., and H.F. planned and designed the study. K.M. collected the data. K.M. and A.M. implemented the neuroimage preprocessing pipeline. K.M., D.G. and L.W. developed and consulted on methods and statistical analyses. K.M. and D.G. drafted the manuscript, and all authors discussed the results, conclusions and edited the manuscript.

