## Supplementary Material 1 for "Moment-to-moment brain signal variability reliably predicts psychiatric treatment outcome"

**This document includes:**

1. Supplementary Methods
2. Supplementary Results
3. Supplementary Figure 1-10
4. Supplementary Tables 1-12

### Supplementary Methods

#### Data diagnostics

The multivariate Mahalanobis distance measure was used to review and identify possible statistical outliers of the prediction model including all treatment outcome predictors (i.e., social anxiety change scores, as predicted by the pre-treatment Liebowitz social anxiety scale, self-report version (LSAS-SR), brain signal variability (task $SD_{BOLD}$, resting-state $SD_{BOLD}$), and average neural response (task $MEAN_{BOLD}$)). As shown in Figure S1, one patient was deemed an outlier and subsequently removed from testing at all levels of analysis in the current study.

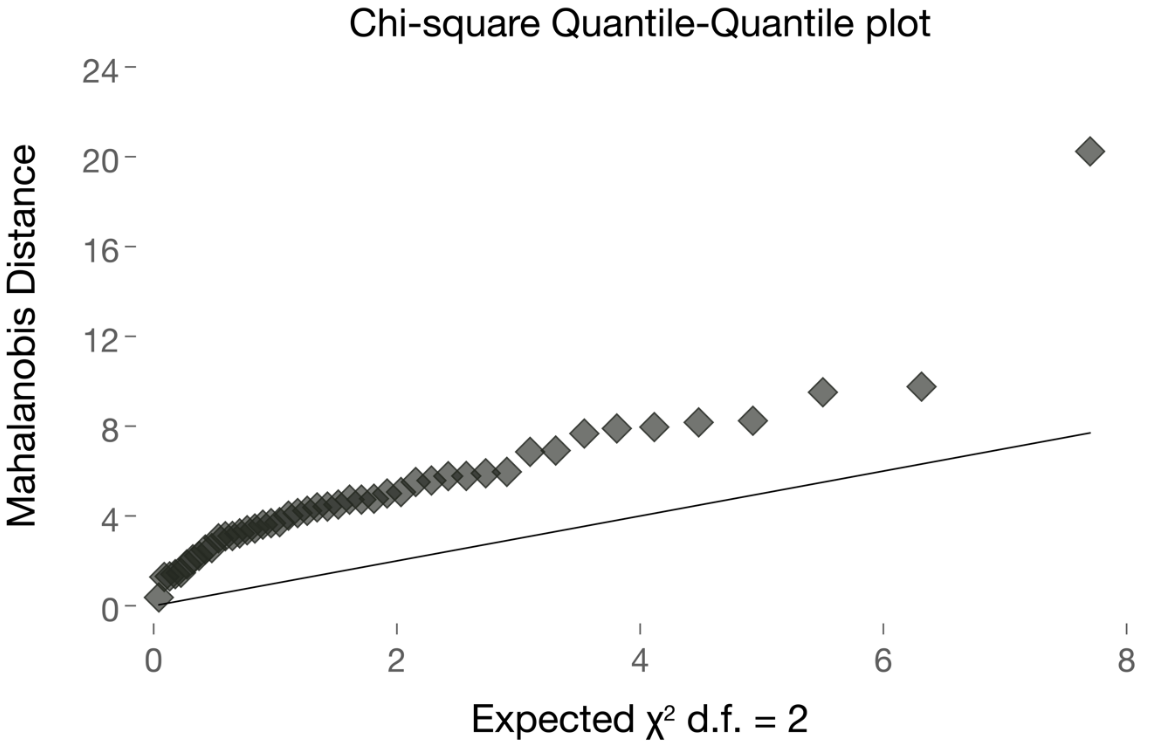


**Figure S1. Detecting outliers.** This figure demonstrates a quantile-to-quantile plot of the Mahalanobis distance measure. The measure is based on all initial predictors (i.e., task $SD_{BOLD}$, resting-state $SD_{BOLD}$, and pre-treatment LSAS-SR) and the LSAS-SR change score as outcome. The model includes all 46 patients initially included in the study and one observation was deemed as a multivariate statistical outlier and thus removed for further testing.

#### Social anxiety disordered patients

Table S1 provides a detailed summary of demographic, clinical status, comorbid mental illness, as well as concurrent medications for patients included in the current study.

**Table S1.** Demographics, clinical status, concurrent psychotropic medications, and comorbid conditions in 45 social anxiety disordered patients.

| **Variable** | **Patients, *n* = 45** | |
|---|---|---|
| Females, *n,* % | 28 | 62.2 |
| Age, average, ±SD | 30.8 | 8.4 |
| SAD duration in years mean, average, ±SD | 17.0 | 10.0 |
| **Marital status, *n*, %** | | |
| Married/cohabiting with children | 16 | 35.6 |
| Married/cohabiting without children | 10 | 22.2 |
| Non-cohabiting partner | 4 | 8.9 |
| Single with children | 3 | 6.7 |
| Single without children | 9 | 20.0 |
| Other | 3 | 6.7 |
| **Education, *n,* %** | | |
| Completed primary school | 3 | 6.7 |
| Completed secondary school | 7 | 15.6 |
| Completed vocational education | 2 | 4.4 |
| Ongoing university education | 15 | 33.3 |
| Completed university education | 18 | 40.0 |
| **Concurrent psychotropic medications, *n,* %** | | |
| No concurrent medication | 41 | 91.1 |
| SSRIs | 4 | 8.9 |
| **Psychiatric comorbidity (M.I.N.I and SCID), *n,* %** | | |
| No concurrent or previous psychiatric comorbidity | 20 | 44.4 |
| Previous depressive episode(s) | 20 | 44.4 |
| Current (and previous) depressive episodes | 1 | 2.2 |
| Current dysthymia | 1 | 2.2 |
| Previous panic disorder | 3 | 6.7 |
| Current panic disorder | 1 | 2.2 |
| Previous agoraphobia | 1 | 2.2 |
| Current generalized anxiety disorder | 1 | 2.2 |

**Abbreviations:** SSRIs, selective serotonin-reuptake inhibitors; M.I.N.I., Mini International Neuropsychiatric Interview; SCID, Structured Clinical Interview for DSM; SAD, Social anxiety disorder;

### Supplementary Results

#### Main and secondary treatment outcomes

As displayed in Table S2, main and secondary social anxiety outcomes, as well as depression and insomnia symptoms decreased significantly over the course of therapy.

**Table S2.** The table includes self-reported main and secondary outcomes from pre- to post-treatment for 45 patients. Group averages and 95% confidence intervals are displayed. Effect sizes represent Cohen's *d* between the first and last assessment of each measure.

| | **Pre-treatment assessments** | | | **Post-treatment assessments** | | |
|---|---|---|---|---|---|---|
| **Self-reports** | **Screening** | **Baseline 1** | **Baseline 2** | **Post-treatment** | **Pre to post change** | |
| | *M* (95% CI) | *M* (95% CI) | *M* (95% CI) | *M* (95% CI) | *M* difference (95% CI) | Effect size (95% CI) |
| **LSAS-SR** | 77.29 (72.2, 82.4) | 75.73 (70.5, 80.9) | 73.33 (67.5, 79.1) | 43.49 (37.1, 49.9) | -33.80 (-39.9, -27.7) | 1.62 (1.2, 2.1) |
| **SIAS** | 55.18 (51.3, 59.1) | - | - | 34.90 (31.1, 38.7) | -20.29 (-24.1, -16.4) | 1.49 (1.1, 1.9) |
| **SPS** | 40.56 (36.4, 44.7) | - | - | 19.60 (16.2, 23.0) | -20.96 (-24.9, -17.0) | 1.57 (1.1, 2.0) |
| **SPSQ** | 34.09 (31.5, 36.7) | - | - | 17.62 (15.1, 20.2) | -16.47 (-19.3, -13.7) | 1.86 (1.3, 2.4) |
| **ISI** | - | 9.22 (7.6, 10.8) | - | 6.98 (5.5, 8.4) | -2.24 (-3.7, -0.8) | 0.42 (0.1, 0.7) |
| **MADRS-S** | 15.62 (14.0, 17.2) | 13.07 (11.2, 15.0) | 12.78 (10.6, 14.9) | 9.04 (6.8, 11.3) | -6.58 (-8.4, -4.8) | 0.95 (0.6, 1.3) |

**Abbreviations:** LSAS-SR, Liebowitz social anxiety scale, self-report version; SIAS, Social interaction anxiety scale; SPS, Social phobia scale; SPSQ, Social phobia screening questionnaire; ISI, Insomnia severity index; MADRS-S, Montgomery-Åsberg depression rating scale, self-report version;

Figure S2A displays the primary measure of social anxiety at each time-point (Screening, Baseline 1, Baseline 2, and Post-treatment), as well as the calculated changes scores (i.e., Post-treatment minus Baseline 1, and Post-treatment minus Baseline 2; Figure S2B). As expected, the two change scores were highly correlated: $r(45) = .86$, $P < 0.001$; Figure S2C).

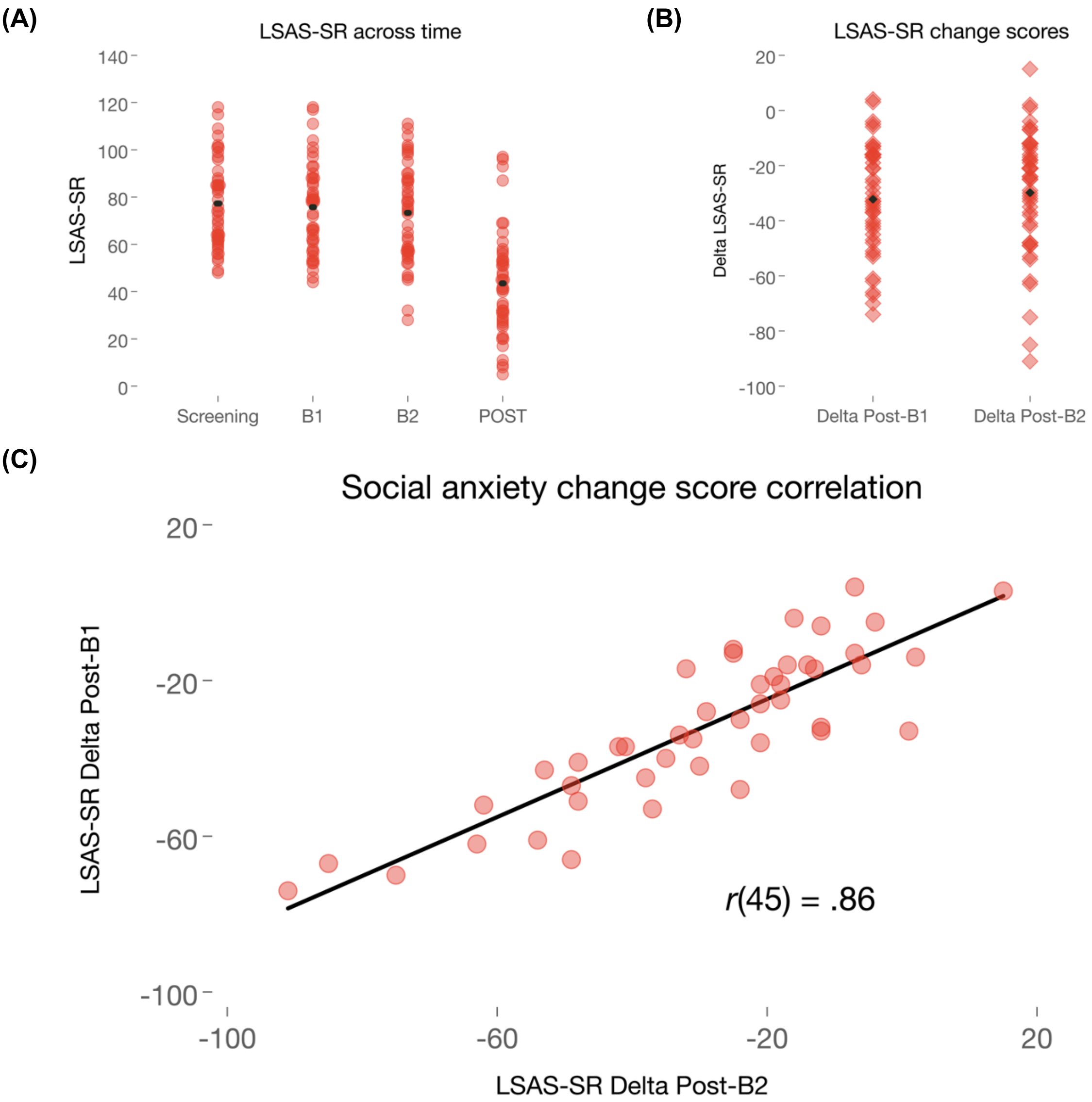


**Figure S2. Main social anxiety treatment outcome.** Figure **(A)** displays the main social anxiety outcome (Liebowitz social anxiety scale, self-report version, LSAS-SR) across all time-points, and **(B)** demonstrates LSAS-SR changes score for each individual and time-period. The black triangles denote the group average scores. **(C)** depicts the correlation between social anxiety change scores (LSAS-SR Post-treatment minus Baseline 1 *versus* LSAS-SR Post-treatment minus Baseline 2). All figures include 45 patients. **Abbreviations:** B1, Baseline 1; B2, Baseline 2; Post, Post-treatment;

Figure S3 displays the secondary self-reported measures of social anxiety and their respective changes scores (i.e., Post-treatment minus Baseline 1). Similarly, the secondary self-reported outcomes (change scores) on depressive and insomnia symptoms are displayed in Figure S3B.

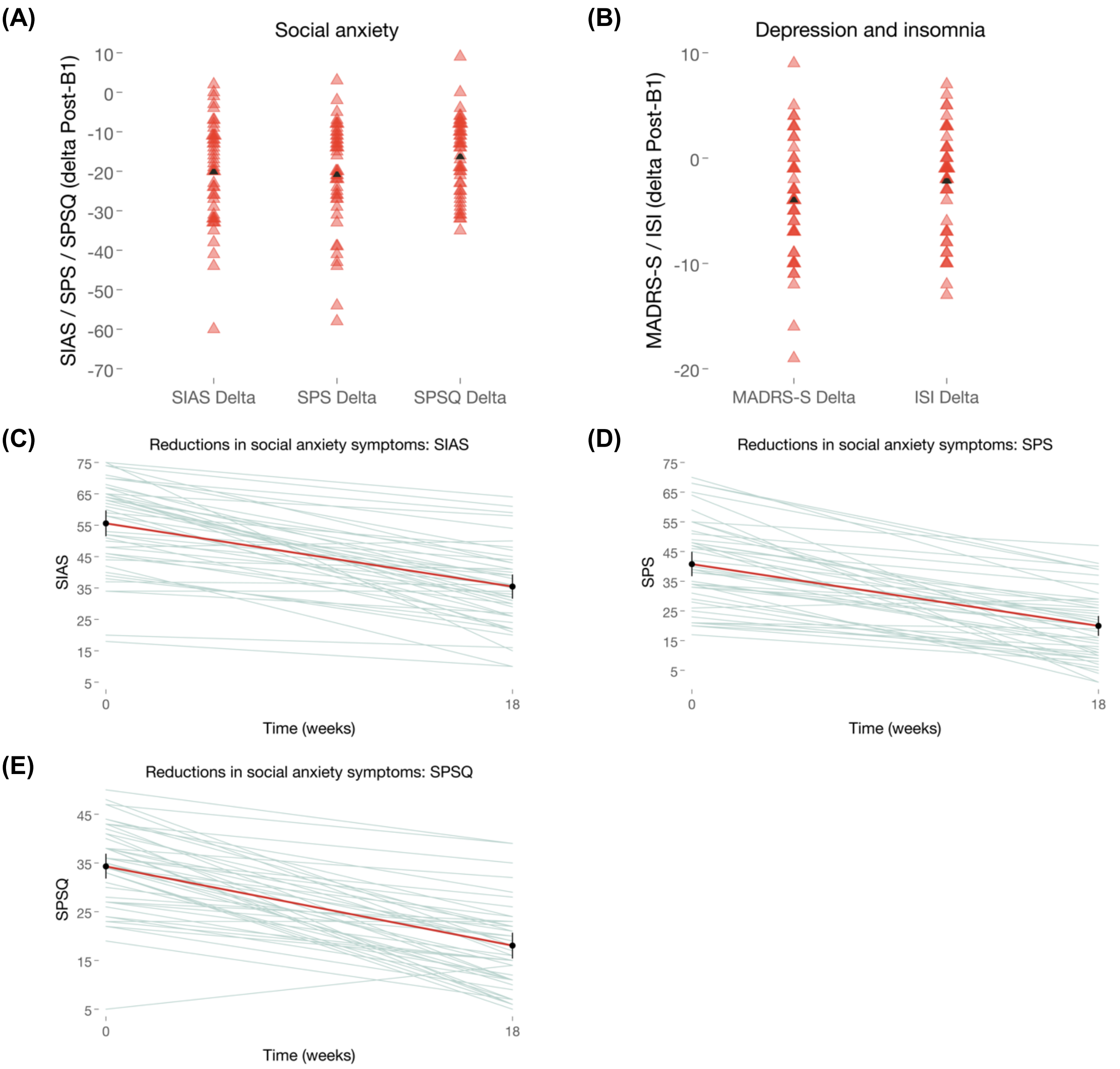


**Figure S3. Secondary treatment outcomes. (A)** displays social anxiety changes scores (i.e., Post-treatment minus Baseline 1) with the black triangle denoting the group average change score, and **(B)** depression and insomnia symptom change score (i.e., Post-treatment minus Baseline 1) with the group's average change score denoted with a black triangle. Also, individual slopes and group averages (solid red line), 95% CI (solid black error bars) are displayed for **(C)** the Social interaction anxiety scale (SIAS), **(D)** the Social phobia scale (SPS), and **(E)** the Social phobia screening questionnaire (SPSQ). All figures include 45 patients. **Abbreviations:** B1, Baseline 1; B2, Baseline 2; Post, Post-treatment;

In addition to all self-reported measures, psychiatric interviews were completed at post-treatment including 45 patients. As displayed in Figure S4, according to the Clinical Global Impression-Improvement (CGI-I) assessment, a majority of the patients showed much or very much improvement after the treatment. Further, remission was determined by use of the DSM-5 criteria, as well as with the M.I.N.I. interview at post-treatment (see Table S3).

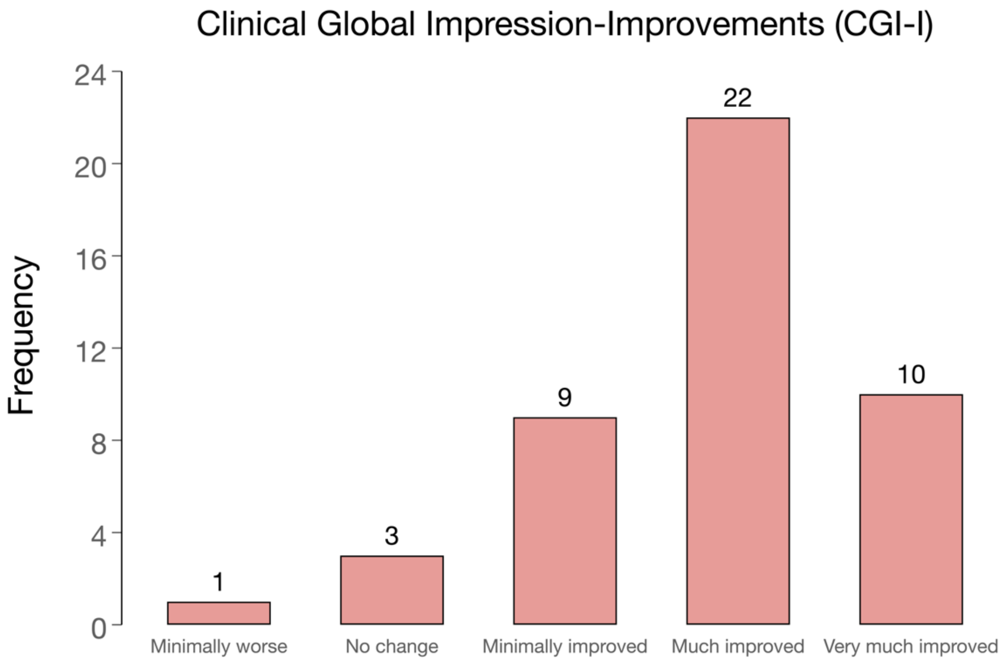


**Figure S4. Clinical Global Impression-Improvements.** The figure displays the clinician administrated CGI-I assessments at post-treatment (*n* = 45). **Abbreviations:** CGI-I, Clinical global impression-improvement;

**Table S3.** Assessment of post-treatment remission status was conducted by use of structured clinical interviews of 45 patients.

| | **M.I.N.I.** | | **DSM-5** | |
|---|---|---|---|---|
| *Indicating SAD diagnosis* | *N* | *%* | *N* | *%* |
| No | 24 | 53.3 | 8 | 17.8 |
| Yes | 21 | 46.7 | 37 | 82.2 |

**Abbreviations:** M.I.N.I., Mini International Neuropsychiatric Interview; DSM-5, The Diagnostic and Statistical Manual of Mental Disorders; SAD, Social anxiety disorder;

#### Treatment outcome predictions

Moment-to-moment brain signal variability during emotional face processing predicted social anxiety change scores with cognitive-behavioral therapy (CBT). As displayed in Table S4, task $SD_{BOLD}$ predicted treatment outcome even after constraining the data volume to 80 or even 40 sec. Table S4 also displays similar predictions and metrics for all other conditions (i.e., self-reported social anxiety, resting-state $SD_{BOLD}$, and task $MEAN_{BOLD}$).

**Table S4.** Zero-order treatment outcome predictions including 45 patients with social anxiety disorder.

| | | | | 95% CI | | |
|---|---|---|---|---|---|---|
| **Pre-treatment predictors** | **Data volume (items)** | **MASE** | ***r*(45)** | **Lower** | **Higher** | **Permuted *P*** |
| **Behavioral, self-reports** | | | | | | |
| LSAS-SR at B1 | 48 | 0.65 | .27 | -.01 | .56 | 0.071 |
| LSAS-SR at B2 | 48 | 0.58 | .45 | .20 | .70 | <0.001 |
| | **Data volume (seconds)** | **MASE** | ***r*(45)** | **Lower** | **Higher** | **Permuted *P*** |
| **Brain, neural response** | | | | | | |
| Task $SD_{BOLD}$ | 160 | 0.54 | .65 | .51 | .79 | <0.001 |
| | 80 | 0.53 | .65 | .52 | .78 | <0.001 |
| | 40 | 0.53 | .62 | .45 | .79 | <0.001 |
| Resting-state $SD_{BOLD}$ | 340 | 0.55 | .55 | .35 | .75 | <0.001 |
| | 160 | 0.63 | .19 | -.08 | .46 | 0.204 |
| | 80 | 0.62 | .24 | -.02 | .51 | 0.112 |
| | 40 | 0.64 | .16 | -.12 | .45 | 0.294 |
| Task $MEAN_{BOLD}$ | 160 | 0.67 | -.18 | -.49 | .12 | 0.218 |
| | 80 | 0.67 | -.28 | -.58 | .03 | 0.084 |
| | 40 | 0.67 | -.20 | -.53 | .12 | 0.179 |

**Abbreviations:** MASE, Mean absolute scaled error; LSAS-SR, Liebowitz social anxiety scale, self-report version; B1, Baseline 1; B2, Baseline 2; $SD_{BOLD}$, Standard deviation of BOLD; $MEAN_{BOLD}$, Average BOLD activity; BOLD, Blood-oxygen-level-dependent imaging;

Behavioral partial least squares (PLS) models on task $SD_{BOLD}$, resting-state $SD_{BOLD}$, and task $MEAN_{BOLD}$ were run to predict the main social anxiety outcome (delta $LSAS\text{-}SR_{POST\text{-}B1}$ and delta $LSAS\text{-}SR_{POST\text{-}B2}$). Baseline 1 and baseline 2 brain scores, and LSAS-SR change scores are displayed in Figure S5.

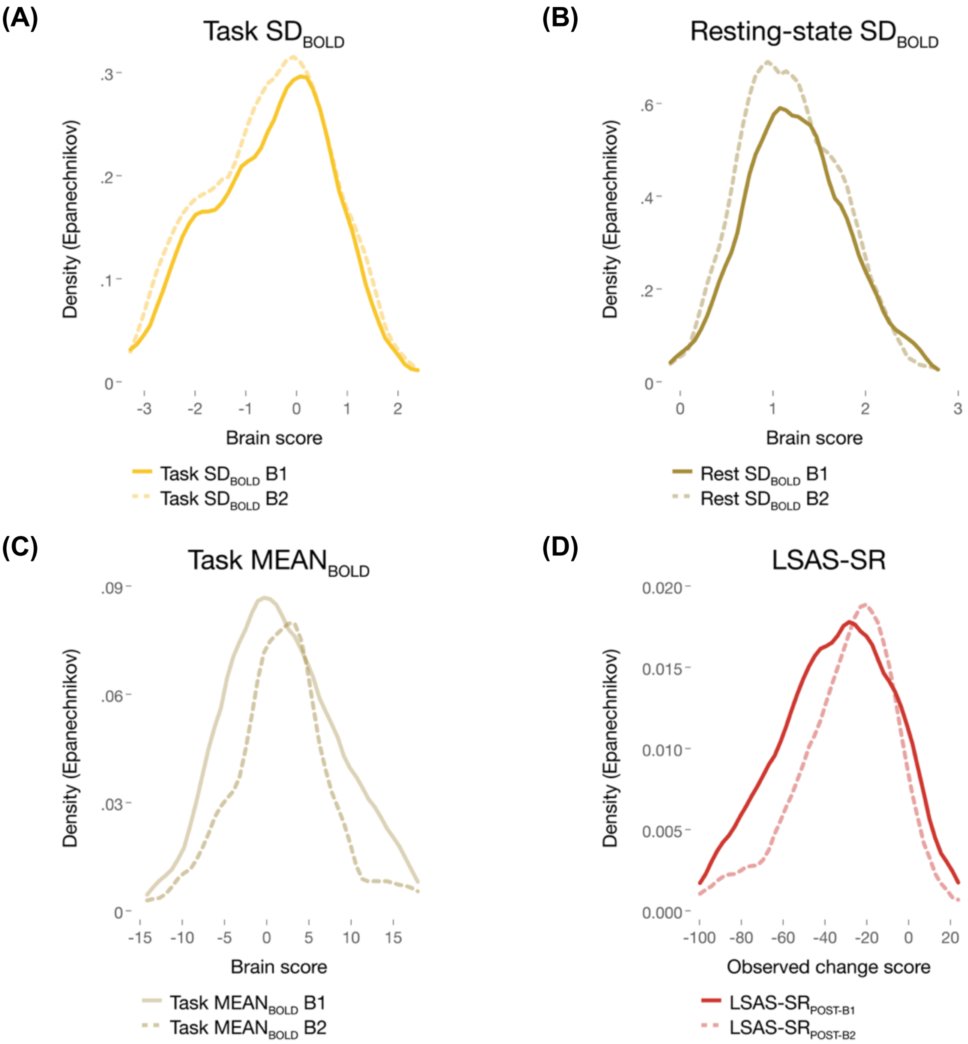


**Figure S5. Data diagnostics.** Behavioral PLS brain scores at the first and second baseline for **(A)** task SD$_{BOLD}$, **(B)** resting-state SD$_{BOLD}$, and **(C)** task MEAN$_{BOLD}$. Each behavioral PLS model includes the LSAS-SR change score (i.e., post-treatment minus baseline) and each model is based on 45 patients. **(D)** displays the LSAS-SR change scores (Post-B1 and Post-B2). **Abbreviations:** PLS, Partial least squares; LSAS-SR, Liebowitz social anxiety scale, self-report version; SD$_{BOLD}$, Standard deviation of BOLD; MEAN$_{BOLD}$, Average BOLD activity; BOLD, Blood-oxygen-level-dependent imaging; B1, Baseline 1; B2, Baseline 2; Post, Post-treatment;

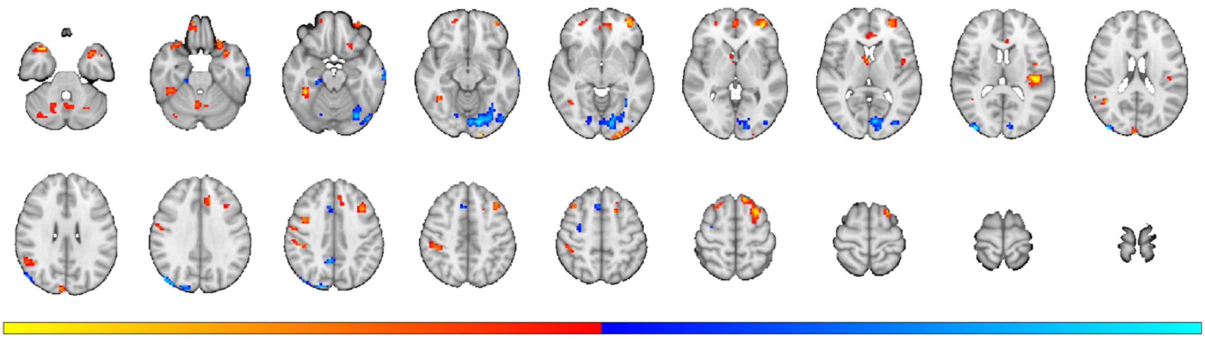

**Figure S6. Task-related SD$_{BOLD}$.** The figure demonstrates task-related variability predicting treatment outcome in 45 patients. Blue regions (negative BSRs) depict less variability, predicting better treatment outcome, whereas red/yellow (positive BSRs) represent high variability predicting better treatment outcome. Neural response pattern is thresholded at BSR ≥ 2, with an extent threshold of 20 voxels, and the minimum distance between clusters is 10 mm. See also Table S6 for details. **Abbreviations:** BSR, Bootstrap ratio; SD$_{BOLD}$, Standard deviation of BOLD; BOLD, Blood-oxygen-level-dependent imaging;

**Table S6. Task-related SD$_{BOLD}$.** The table depicts task-related variability predicting treatment outcome in 45 patients. Negative BSRs depict less variability, predicting better treatment outcome, whereas positive BSRs represent high variability predicting better treatment outcome. Neural response pattern is thresholded at BSR = 2, with an extent threshold of 20 voxels, and the minimum distance between clusters is 10 mm.

| **Behavioral PLS** | **MNI coordinates** | | | | |
|---|---|---|---|---|---|
| **Region** | **x** | **y** | **z** | **BSR** | **k, voxels** |
| Middle Occipital Gyrus | -39 | -90 | 18 | -4.8833 | 54 |
| Lingual Gyrus | 15 | -81 | -12 | -4.5411 | 453 |
| ParaHippocampal Gyrus | -21 | -27 | -21 | -4.5362 | 20 |
| Superior Occipital Gyrus | -15 | -90 | 36 | -4.2837 | 92 |
| Precuneus | -6 | -51 | 39 | -3.3357 | 24 |
| Middle Temporal Gyrus | 66 | -18 | -21 | -3.3318 | 40 |
| Posterior-Medial Frontal | -6 | 21 | 48 | -3.1836 | 38 |
| Lingual Gyrus | -15 | -72 | -9 | -2.7278 | 23 |
| Precentral Gyrus | -33 | -6 | 57 | -2.5941 | 21 |
| Heschls Gyrus | 45 | -24 | 15 | 5.5041 | 96 |
| Middle Orbital Gyrus | 36 | 48 | -3 | 5.3999 | 111 |
| Inferior Temporal Gyrus | -39 | -39 | -18 | 4.7366 | 79 |
| Temporal Pole | 33 | 12 | -27 | 4.5093 | 45 |
| Temporal Pole | -33 | 18 | -30 | 4.3716 | 90 |
| Lingual Gyrus | 18 | -99 | -9 | 4.3196 | 36 |
| Superior Frontal Gyrus | 27 | 18 | 63 | 4.2674 | 130 |
| Angular Gyrus | -42 | -54 | 24 | 4.1736 | 47 |
| Medial Temporal Pole | 45 | 9 | -36 | 3.6766 | 25 |
| Rolandic Operculum | 45 | -3 | 12 | 3.5904 | 21 |
| Mid Cingulate Cortex | 12 | 33 | 30 | 3.4495 | 45 |
| Cerebelum VI | 27 | -60 | -33 | 3.4281 | 104 |
| Middle Frontal Gyrus | 39 | 18 | 39 | 3.3858 | 62 |
| Middle Frontal Gyrus | -33 | 21 | 57 | 3.1685 | 35 |
| Thalamus | -3 | 0 | 3 | 3.1629 | 21 |
| Cuneus | -6 | -90 | 24 | 3.1627 | 32 |
| Inferior Frontal Gyrus | 24 | 18 | -21 | 3.0661 | 36 |
| Mid Orbital Gyrus | 0 | 51 | -3 | 3.0557 | 34 |

| | | | | | |
|---|---|---|---|---|---|
| Precentral Gyrus | -39 | 3 | 39 | 3.0385 | 29 |
| Postcentral Gyrus | -42 | -33 | 42 | -3.0383 | 68 |
| Postcentral Gyrus | -57 | -6 | 36 | -2.9871 | 21 |
| Superior Orbital Gyrus | -12 | 54 | -24 | -2.9376 | 25 |
| Superior Orbital Gyrus | -27 | 51 | -3 | -2.8913 | 23 |
| Cerebelum Crus 1 | -36 | -72 | -27 | -2.8403 | 21 |
| Cerebellar Vermis 8 | 0 | -66 | -42 | -2.7868 | 66 |
| Anterior Cingulate Cortex | 0 | 30 | 12 | -2.6862 | 30 |
| Dorsal Dentate Nucleus | -18 | -60 | -33 | -2.6797 | 55 |

**Abbreviations:** BSR, Bootstrap ratio; $SD_{BOLD}$, Standard deviation of BOLD; BOLD, Blood-oxygen-level-dependent imaging;

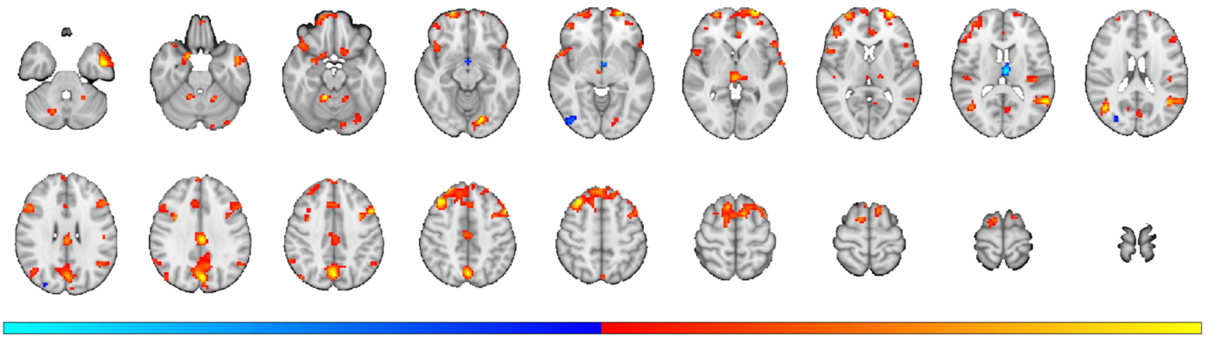

**Figure S7. Task-related $MEAN_{BOLD}$.** The figure demonstrates task-related mean neural response predicting treatment outcome in 45 patients. Blue regions (negative BSRs) depict less activity, predicting better treatment outcome, whereas yellow (positive BSRs) represent more activity predicting better treatment outcome. Neural response pattern is thresholded at BSR ≥ 2, with an extent threshold of 20 voxels, and the minimum distance between clusters is 10 mm. See also Table S7 for details. **Abbreviations:** BSR, Bootstrap ratio; $MEAN_{BOLD}$, Average BOLD activity; BOLD, Blood-oxygen-level-dependent imaging;

**Table S7. Task-related $MEAN_{BOLD}$.** The table depicts task-related mean neural response predicting treatment outcome in 45 patients. Negative BSRs depict less activity, predicting better treatment outcome, whereas positive BSRs represent more activity predicting better treatment outcome. Neural response pattern is thresholded at BSR = 2, with an extent threshold of 20 voxels, and the minimum distance between clusters is 10 mm.

| **Behavioral PLS** | **MNI coordinates** | | | | |
|---|---|---|---|---|---|
| **Region** | **x** | **y** | **z** | **BSR** | **k, voxels** |
| Middle Temporal Gyrus | 51 | -3 | -30 | 4.9126 | 154 |
| Superior Orbital Gyrus | 27 | 66 | -3 | 4.8444 | 193 |
| Middle Frontal Gyrus | 51 | 15 | 42 | 4.4551 | 871 |
| Cuneus | 3 | -75 | 33 | 4.4536 | 448 |
| Middle Frontal Gyrus | -33 | 27 | 51 | 4.4255 | 211 |
| Cerebelum IV-V | -12 | -48 | -21 | 4.199 | 39 |
| Lingual Gyrus | 18 | -78 | -12 | 4.186 | 117 |
| Middle Temporal Gyrus | -42 | -63 | 21 | 4.1779 | 111 |
| Mid Cingulate Cortex | 3 | -24 | 33 | 4.0787 | 155 |
| Middle Temporal Gyrus | 57 | -54 | 15 | 4.0547 | 182 |
| Middle Frontal Gyrus | -27 | 63 | 3 | 3.8269 | 351 |

| | | | | | |
|---|---|---|---|---|---|
| Thalamus | -3 | -24 | 0 | 3.6117 | 82 |
| ParaHippocampal Gyrus | -18 | 9 | -24 | 3.6114 | 55 |
| Superior Temporal Gyrus | 63 | -3 | 3 | 3.4839 | 37 |
| Precentral Gyrus | -33 | 6 | 33 | 3.4639 | 31 |
| Inferior Frontal Gyrus | -54 | 21 | 30 | 3.4046 | 59 |
| Cerebelum IV-V | 18 | -51 | -24 | 3.366 | 34 |
| Cerebelum Crus 2 | 6 | -72 | -33 | 3.1282 | 36 |
| SupraMarginal Gyrus | -63 | -51 | 36 | 3.0459 | 26 |
| Cerebelum VIII | 24 | -57 | -48 | 3.0056 | 35 |
| Temporal Pole | -45 | 24 | -15 | 2.9605 | 134 |
| Anterior Cingulate Cortex | 6 | 39 | 0 | 2.9443 | 46 |
| Superior Temporal Gyrus | -51 | -21 | 12 | 2.9397 | 23 |
| Cerebelum Crus 1 | -21 | -66 | -33 | 2.9329 | 28 |
| Cerebelum Crus 2 | -39 | -66 | -39 | 2.8828 | 56 |
| Heschls Gyrus | 42 | -24 | 15 | 2.8633 | 53 |
| Superior Medial Gyrus | -3 | 57 | 39 | 2.7485 | 27 |
| Inferior Frontal Gyrus | 54 | 21 | 0 | 2.7409 | 40 |
| Olfactory cortex | 15 | 12 | -18 | 2.6697 | 20 |
| Middle Frontal Gyrus | 33 | 51 | 0 | 2.4683 | 26 |
| Thalamus | 3 | -12 | 15 | -4.1941 | 44 |
| Thalamus | 3 | -3 | -9 | -3.6736 | 20 |
| Inferior Occipital Gyrus | -42 | -81 | -6 | -2.7712 | 27 |
| Middle Occipital Gyrus | -30 | -78 | 21 | -2.5005 | 20 |

**Abbreviations:** BSR, Bootstrap ratio; $MEAN_{BOLD}$, Average BOLD activity; BOLD, Blood-oxygen-level-dependent imaging;

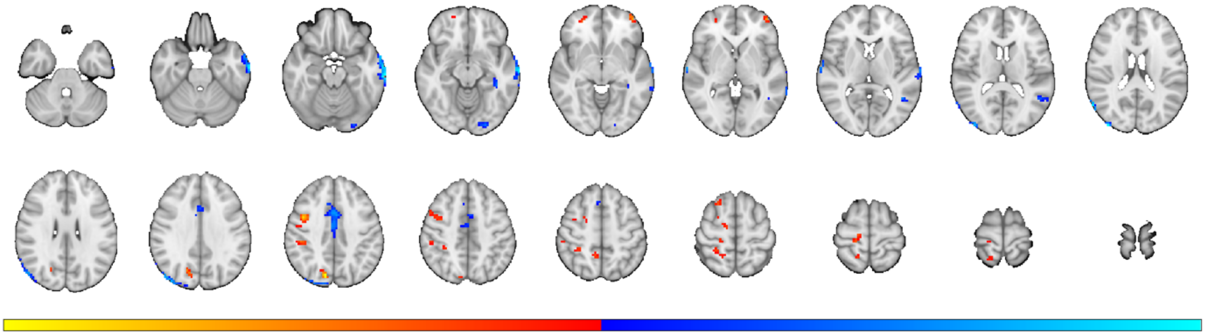

**Figure S8. Resting-state $SD_{BOLD}$.** The figure demonstrates resting-state neural variability predicting treatment outcome in 45 patients. Blue regions (negative BSRs) depict less variability, predicting better treatment outcome, whereas red/yellow (positive BSRs) represent high variability predicting better treatment outcome. Neural response pattern is thresholded at BSR ≥ 2, with an extent threshold of 20 voxels, and the minimum distance between clusters is 10 mm. See also Table S8 for details. **Abbreviations:** BSR, Bootstrap ratio; $SD_{BOLD}$, Standard deviation of BOLD; BOLD, Blood-oxygen-level-dependent imaging;

**Table S8. Resting-state $SD_{BOLD}$.** The table depicts resting-state variability predicting treatment outcome in 45 patients. Negative BSRs depict less variability, predicting better treatment outcome, whereas positive BSRs represent high variability predicting better treatment outcome. Neural response pattern is thresholded at BSR = 2, with an extent threshold of 20 voxels, and the minimum distance between clusters is 10 mm.

| **Behavioral PLS** | **MNI coordinates** | | | | |
|---|---|---|---|---|---|
| **Region** | **x** | **y** | **z** | **BSR** | **k, voxels** |
| Middle Temporal Gyrus | 69 | -21 | -18 | -4.2981 | 199 |
| Middle Occipital Gyrus | -39 | -90 | 18 | -4.2196 | 142 |
| Mid Cingulate Cortex | 0 | -6 | 42 | -4.1964 | 61 |
| Mid Cingulate Cortex | 3 | 6 | 42 | -3.2932 | 92 |
| Middle Temporal Gyrus | -66 | -12 | 0 | -3.2433 | 26 |
| Lingual Gyrus | 27 | -90 | -15 | -3.2080 | 32 |
| Middle Temporal Gyrus | 45 | -54 | 3 | -3.1586 | 40 |
| Fusiform Gyrus | 39 | -36 | -12 | -2.8847 | 23 |
| Cuneus | -12 | -75 | 39 | 4.0101 | 45 |
| Middle Orbital Gyrus | 42 | 54 | -3 | 3.6862 | 24 |
| Precentral Gyrus | -42 | 3 | 39 | 3.6293 | 72 |
| Postcentral Gyrus | -18 | -27 | 63 | 2.8923 | 27 |
| Inferior Parietal Lobule | -45 | -30 | 39 | 2.8885 | 113 |
| Superior Orbital Gyrus | -27 | 51 | -3 | 2.7853 | 20 |
| Middle Frontal Gyrus | -24 | 21 | 60 | 2.6437 | 21 |
| Middle Frontal Gyrus | -18 | -6 | 57 | 2.4842 | 24 |

**Abbreviations:** BSR, Bootstrap ratio; $SD_{BOLD}$, Standard deviation of BOLD; BOLD, Blood-oxygen-level-dependent imaging;

In Table S9, a multiple regression model is presented and includes all predictors of treatment outcome (i.e., actual LSAS-SR change score). This model includes all brain predictors when data volume remains constant across all conditions (i.e., 160 seconds stimuli duration), and shows that task-related neural variability outperforms all other potential predictors of treatment outcome. In a similar vein, Table S10 demonstrates a multiple regression model including all predictors of treatment outcome (i.e., actual LSAS-SR change score). This model includes unequal data volumes for the brain predictors. Specifically, task $SD_{BOLD}$ and $MEAN_{BOLD}$ includes 160 seconds respectively, whereas the resting-state condition includes 340 seconds data. The model shows that the relatively shorter task-related neural variability remains the strongest treatment outcome predictor, and that the longer resting-state condition significantly contributed with some unique variance, see also Figure 4 in the main manuscript for details.

**Table S9.** Multiple prediction model including all fMRI conditions and self-reported social anxiety. Here, all conditions included equal data volumes (i.e., 160 sec). The regression includes 45 patients and the model's adjusted $R^2$ was 54%.

| Predictors | Data volume (seconds) | ***β*** | 95% CI Lower | 95% CI Higher | ***Z*** | Permuted ***P*** |
|---|---|---|---|---|---|---|
| LSAS-SR at B2 | - | .22 | -.02 | .46 | 1.81 | 0.186 |
| Task $SD_{BOLD}$ | 160 | .61 | .41 | .82 | 5.85 | <0.001 |
| Resting-state $SD_{BOLD}$ | 160 | .26 | .07 | .46 | 2.63 | 0.090 |
| Task $MEAN_{BOLD}$ | 160 | -.07 | -.32 | .17 | 0.60 | 0.621 |

**Abbreviations:** fMRI, functional magnetic resonance imaging; $SD_{BOLD}$, Standard deviation of BOLD; $MEAN_{BOLD}$, Average BOLD activity; BOLD, Blood-oxygen-level-dependent imaging; Rest, Resting-state; B2, Baseline 2;

**Table S10.** Multiple prediction model including all fMRI conditions and self-reported social anxiety. Here, task-related fMRI conditions included 160 sec of data volumes, whereas the resting-state condition included 340 sec. The regression includes 45 patients and the model's adjusted $R^2$ was 57%.

| Predictors | Data volume (seconds) | ***β*** | 95% CI Lower | 95% CI Higher | ***Z*** | Permuted ***P*** |
|---|---|---|---|---|---|---|
| LSAS-SR at B2 | - | .29 | .08 | .50 | 2.65 | 0.071 |
| Task $SD_{BOLD}$ | 160 | .41 | .20 | .62 | 3.83 | 0.018 |
| Resting-state $SD_{BOLD}$ | 340 | .34 | .09 | .58 | 2.68 | 0.039 |
| Tast $MEAN_{BOLD}$ | 160 | -.08 | -.28 | .12 | 0.78 | 0.631 |

**Abbreviations:** fMRI, functional magnetic resonance imaging; $SD_{BOLD}$, Standard deviation of BOLD; $MEAN_{BOLD}$, Average BOLD activity; BOLD, Blood-oxygen-level-dependent imaging; Rest, Resting-state; B2, Baseline 2;

As reported in the main manuscript, we computed PLS-based neurobehavioral correlations between the first baseline blood-oxygen-level-dependent (BOLD) activity and reductions in LSAS-SR scores, and the resulting BSRs were thresholded at $\pm$ 2 ($P$ < 0.05) while excluding all clusters smaller than 20 voxels. These thresholded maps were used to extract weights, and applied to MR data recorded at the second baseline measurement (11 weeks after the first baseline) to extract subject-specific brain scores. In addition to this procedure, here we also demonstrate model performance with variable BSR thresholding. Figure S9 A) displays Pearson's *r*, B) mean absolute scaled error (MASE), and C) intraclass correlation coefficient (ICC) for BSR ratios at 0 (i.e., no threshold applied), 1, 2, and 3 for the main condition: task-related $SD_{BOLD}$. In general, Figure S9 demonstrates that predictive and reliability performance peaks at BSR level at 2, and deteriorates at an even stricter level. Interestingly, in a multiple prediction framework (i.e., including all potential pre-treatment brain predictors (task and resting-state $SD_{BOLD}$) and self-reported social anxiety), the prediction performance remains stable and accurate even without applying a threshold based on data from the first baseline assessment (Pearson's $r_{ACT,PRED}$ = .69, MASE = 0.47, $P$ = 0.001; see also Figure S9 D).

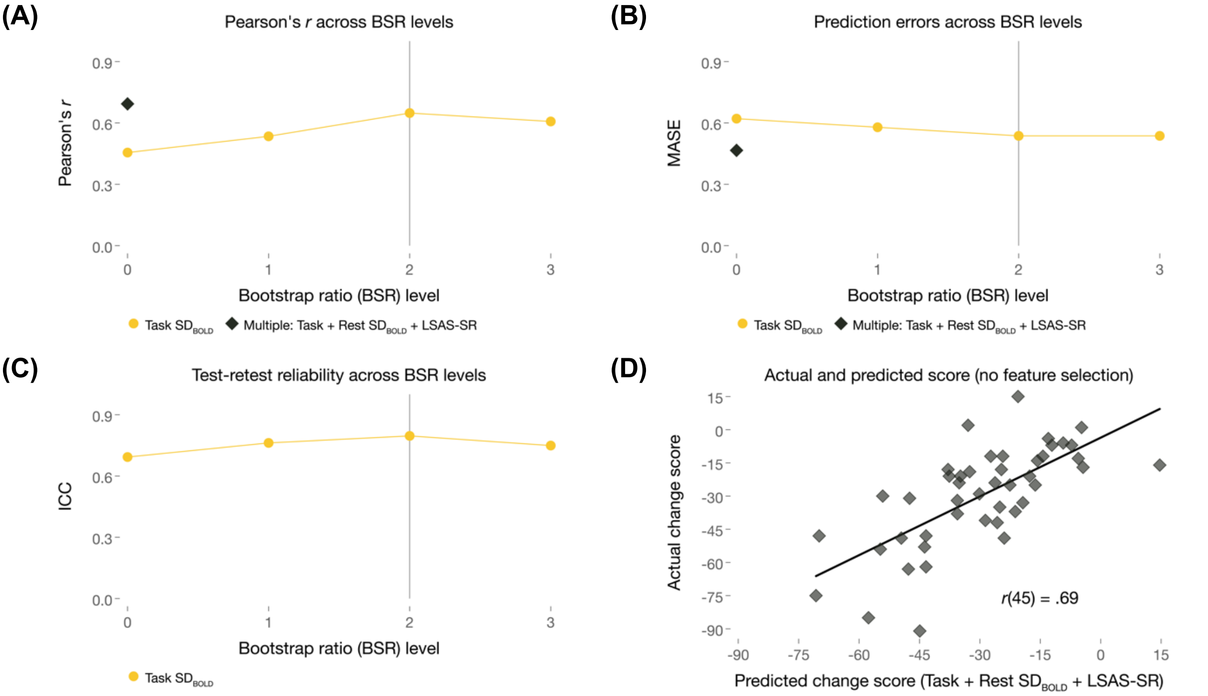


**Figure S9. Prediction accuracies and test-retest reliabilities. (A)** depicts Pearson's $r_{ACTUAL,PREDICTED}$, **(B)** MASE, and **(C)** ICC across differently thresholded behavioral PLS models based on task-related variability (task $SD_{BOLD}$). The multiple prediction model (denoted with black diamonds) is based on the zero-order variables found to significantly predict treatment outcome (i.e., 160 sec task $SD_{BOLD}$, 340 sec resting-state $SD_{BOLD}$, and self-reported pre-treatment social anxiety at the second baseline), and in this forward prediction model a threshold was not set (i.e., no "feature selection") based on the outcome from the first baseline (see detailed description in the main manuscript). **(D)** demonstrates the correlation between the actual social anxiety change score, and the predicted social anxiety change score based on multiple predictors (i.e., task $SD_{BOLD}$, resting-state $SD_{BOLD}$, and pre-treatment LSAS-SR). **Abbreviations:** MASE, Mean absolute scaled error; ICC, Intraclass correlation coefficient; BSR, Bootstrap ratio; PLS, Partial least squares; LSAS-SR, Liebowitz social anxiety scale, self-report version; $SD_{BOLD}$, Standard deviation of BOLD; BOLD, Blood-oxygen-level-dependent imaging;

#### Unique and shared neural variability between the socio-affective task and resting-state fMRI

Figure S10 displays the unique and overlapping brain regions comparing $SD_{BOLD}$ for task and resting-state. Specifically, purple color depicts overlapping task- and resting-state related variability predicting treatment outcome. Blue color depicts unique variability within the task condition, and yellow color depicts unique variability as determined by the longer resting-state condition. In conclusion, the two conditions are largely independent and non-overlapping.

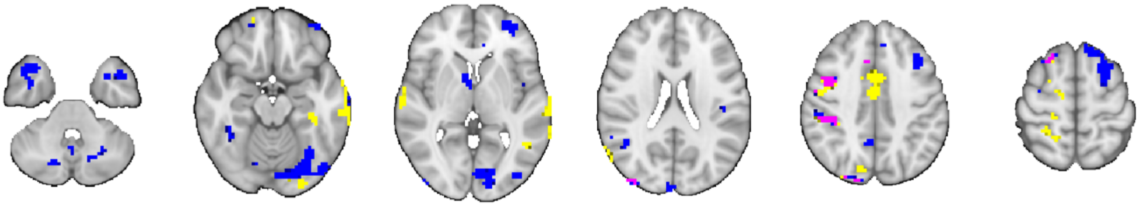

**Figure S10. Task and resting-state $SD_{BOLD}$ overlap.** The figure demonstrates overlapping (purple) and unique neural activity for $SD_{BOLD}$ task (160 sec condition; blue) and $SD_{BOLD}$ resting-state (340 sec condition; yellow) predicting treatment outcome. The spatial pattern for each condition represents a binary mask and thus, direction (low/high variability) cannot be inferred. **Abbreviations:** $SD_{BOLD}$, standard deviation of BOLD; BOLD, Blood-oxygen-level-dependent imaging;

### Test-retest reliability

The 11-week test-retest reliability (Baseline 1 vs Baseline 2) was excellent for the task-related $SD_{BOLD}$ prediction model, and the reliability improved as a function of data volume. Specifically, the intraclass correlation coefficient ($ICC_{B1,B2}$) was lower for the shortest $SD_{BOLD}$ condition (40 sec), and reliability was larger when 160 sec stimuli was included, see details in Table S11. In contrast, task-related $MEAN_{BOLD}$ showed poor reliability (all ICC’$s_{B1,B2}$ ~0) across all conditions. The longer (340 sec) resting-state condition showed excellent test-retest reliability, comparable to the shorter task-related $SD_{BOLD}$ condition, whereas the shorter resting-state conditions only showed poor to good reliability.

**Table S11.** Test-retest reliability of the brain scores included in each treatment outcome prediction model. Each ICC computation included 45 patients.

| **Predictor** | **Data volume (seconds)** | **ICC** | **95% CI (bias corrected) Lower** | **Upper** |
|---|---|---|---|---|
| Task $SD_{BOLD}$ | 40 | 0.62 | 0.44 | 0.78 |
| | 80 | 0.78 | 0.64 | 0.87 |
| | 160 | 0.80 | 0.68 | 0.87 |
| Resting-state $SD_{BOLD}$ | 40 | 0.36 | 0.08 | 0.58 |
| | 80 | 0.56 | 0.31 | 0.73 |
| | 160 | 0.67 | 0.52 | 0.80 |
| | 340 | 0.81 | 0.74 | 0.88 |
| Task $MEAN_{BOLD}$ | 40 | -0.15 | -0.38 | 0.15 |
| | 80 | -0.12 | -0.36 | 0.17 |
| | 160 | -0.05 | -0.35 | 0.29 |

**Abbreviations:** ICC, Intraclass correlation coefficient; BOLD, Blood-oxygen level-dependent; LB, Lower bound confidence intervals; Avg, Average; $SD_{BOLD}$, Standard deviation of BOLD; $MEAN_{BOLD}$, Average BOLD activity; BOLD, Blood-oxygen-level-dependent imaging;

In addition to the reliability of the prediction models, voxel-wise ICC's are displayed in Table S12. In contrast to the previously presented reliability of the treatment outcome predictions, here, all voxels across the whole brain were included from each time-point (Baseline 1 and Baseline 2). Thus, these models represent a global measure of reliability across the whole-brain, rather than reliability of the condition (i.e., task or resting-state), or the treatment outcome prediction model.

**Table S12.** Test-retest reliability across all voxels in the whole-brain ($k$ = 51 609). Average ICCs across 45 patients' whole-brain voxels are displayed below.

| Predictor | Data volume (seconds) | Avg ICC | 95% CI Lower | 95% CI Upper |
|---|---|---|---|---|
| Task $SD_{BOLD}$ | 40 | 0.38 | 0.11 | 0.60 |
| | 80 | 0.46 | 0.20 | 0.65 |
| | 160 | 0.53 | 0.30 | 0.71 |
| Resting-state $SD_{BOLD}$ | 40 | 0.35 | 0.08 | 0.58 |
| | 80 | 0.48 | 0.23 | 0.67 |
| | 160 | 0.58 | 0.36 | 0.74 |
| | 340 | 0.62 | 0.41 | 0.77 |
| Task $MEAN_{BOLD}$ | 40 | 0.03 | -0.26 | 0.31 |
| | 80 | 0.03 | -0.25 | 0.31 |
| | 160 | 0.08 | -0.21 | 0.35 |

**Abbreviations:** ICC, Intraclass correlation coefficient; LB, Lower bound confidence intervals; Avg, Average; $SD_{BOLD}$, Standard deviation of BOLD; $MEAN_{BOLD}$, Average BOLD activity; BOLD, Blood-oxygen-level-dependent imaging;
